# MIMIC-IV Phenotype Atlas (MIPA): A Publicly Available Dataset for EHR Phenotyping

**DOI:** 10.64898/2026.04.16.26350888

**Authors:** Eric Yamga, Reda Goudrar, Philippe Després

**Affiliations:** Department of Medicine, Centre hospitalier de l’Université de Montréal, Montréal, Québec, Canada; Centre de recherche du Centre hospitalier de l’Université de Montréal (CRCHUM), Montréal, Québec, Canada; Université de Montréal; Département de physique, de génie physique et d’optique, Université Laval, Québec, Québec, Canada; Centre de recherche de l’Institut universitaire de cardiologie et de pneumologie de Québec-Université Laval, Québec, Québec, Canada

**Keywords:** Electronic health records, phenotyping, MIMIC-IV, benchmarking dataset, machine learning, large language models, GPT-4o, ICD codes, TF-IDF, supervised learning

## Abstract

Secondary use of electronic health records (EHRs) often requires transforming raw clinical information into research-grade data. A central step in this process is EHR phenotyping - the identification of patient cohorts defined by specific medical conditions. Although numerous approaches exist, from ICD-based heuristics to supervised learning and large language models (LLMs), the field lacks standardized benchmark datasets, limiting reproducibility and hindering fair comparison across methods.

**Method:** We developed the MIMIC-IV Phenotype Atlas (MIPA) dataset, an adaptation of MIMIC-IV that provides expert-annotated discharge summaries across 16 phenotypes of varying prevalence and complexity. Two independent clinicians reviewed and labeled the discharge summaries, resolving disagreements by consensus. In parallel, we implemented a processing pipeline that extracts multimodal EHR features and generates training, validation, and testing datasets for supervised phenotyping.

To illustrate MIPA’s utility, we benchmarked four phenotyping methods: ICD-based classifiers, keyword-driven Term Frequency-Inverse Document Frequency (TF-IDF) classifiers, supervised machine learning (ML) models, and LLMs on the task.

**Results:** The final MIPA corpus consists of 1,388 expert-annotated discharge summaries. Annotation reliability was high (mean document-level kappa = 0.805, mean label-level kappa = 0.771), with 91% of disagreements resolved through consensus review. MIPA provides high-quality phenotype labels paired with structured EHR features and predefined train/validation/test splits for each phenotype.

In the benchmarking case study, LLMs achieved the highest F1 scores in 13 of 16 phenotypes, particularly for conditions requiring contextual interpretation of clinical narrative, while supervised ML offered moderate improvements over rule-based baselines.

**Conclusion:** MIPA is the first publicly available benchmark dataset dedicated to EHR phenotyping, combining expert-curated annotations, broad phenotype coverage, and a reproducible processing pipeline. By enabling standardized comparison across ICD-based heuristics, ML models, and LLMs, MIPA provides a durable reference resource to advance methodological development in automated phenotyping.

## 1.1 Introduction

Electronic health records (EHRs) are a rich source of data for clinical research, supporting applications ranging from epidemiologic studies to disease prediction models^1,2^. A critical prerequisite for secondary use of EHR data is phenotyping— cohort identification of patients with a clinical condition of interest ^3^. Over the past decade, computational approaches have evolved from rule-based systems to machine learning models and, more recently, large language models capable of processing unstructured clinical text^4–9^.

Despite these advances, progress in EHR phenotyping has been hindered by the absence of standardized, openly available benchmark datasets. Most published algorithms are evaluated on institution-specific data with heterogeneous case definitions and locally curated labels, which limits reproducibility and prevents fair head-to-head comparison across methods^2,4^. As a result, the field lacks a common foundation for systematically assessing different approaches to computational phenotyping^10^. We therefore developed the MIMIC-IV Phenotype Atlas (MIPA), a clinician-annotated dataset specifically designed to support systematic benchmarking of EHR phenotyping approaches. In this paper, we describe the dataset construction process and demonstrate its utility by evaluating various methods on the dataset.

Although a number of publicly available EHR related datasets exist, none are specifically tailored for computational phenotyping. Notably, there are clinical natural language processing (NLP) datasets that consist of annotated clinical notes without their corresponding EHR structured counterpart. Notably, the n2c2 shared tasks have released corpora of deidentified discharge summaries annotated for smoking status (n2c2 2006, n=502)^11^, obesity related health conditions (n2c2 2008 n=1237)^12^ and more recently, n2c2 2018 n= 288, a dataset specifically annotated to benchmark clinical trial patient matching^13^, a task closely related to computational phenotyping. On the other hand, there are publicly available comprehensive EHR datasets that contain longitudinal structured data but not clinical notes and are thus not tailored for cohort identification related tasks^14,15^.

To address this gap, we developed the MIMIC-IV Phenotype Atlas (MIPA), the first publicly available, annotated dataset specifically designed for benchmarking EHR phenotyping. Derived from MIMIC-IV^16^, MIPA encompasses both structured longitudinal EHR data as well as de-identified clinical notes in the form of radiology reports and discharge summaries (see Figure 1). Beyond its immediate utility for training and evaluating phenotyping models, MIPA serves as a shared reference point for reproducible research and assessment of phenotyping algorithms.

**Figure 1.**
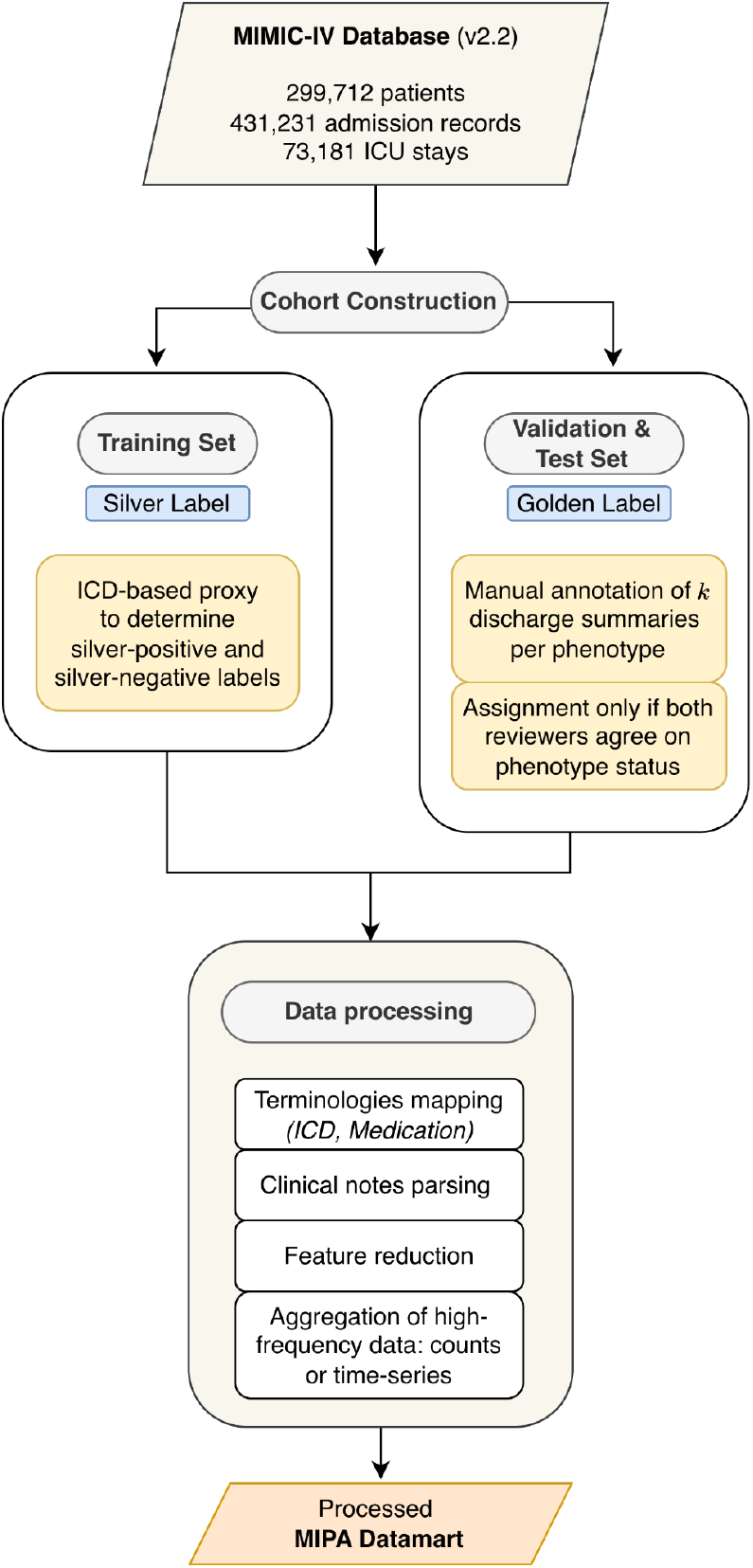
MIPA phenotyping benchmark database development pipeline

## 1.2 Material and Methods

### 1.2.1 Dataset

To construct MIPA, we leveraged the publicly available MIMIC-IV 2.2 database (see Fig. 1). This database contains admission records for adult patients at the Beth Israel Deaconess Medical Center between 2008 and 2019, including 431,231 hospital admissions for 180,733 unique patients and 73,181 intensive care unit (ICU) admissions for 50,920 unique patients. It includes both structured medical data and de-identified clinical notes, anonymized according to the HIPAA (Health Insurance Portability and Accountability Act) Safe Harbor standards. Information removed includes patient social history, discharge instructions, and any other identifiers that could reveal patient identity. Researchers can ask for authorized access to MIMIC-IV through PhysioNet, adhering to all data use agreements^16,17^.

### 1.2.2 Phenotypes and Annotation

In designing our annotation and consensus process, we sought to align with principles promoted by the ACCORD (ACcurate COnsensus Reporting Document) guideline for transparent reporting of consensus-based methods^18^. First, we identified sixteen phenotypes for the dataset reflecting conditions of varying prevalence, complexity and temporality (see Table 1). For each phenotype, we curated expert-defined ICD code lists considered indicative of the condition (see Supplementary File 1). We then identified a set of approximately 100 candidate discharge summaries per phenotype based on the presence of at least one ICD-code related to the phenotype, yielding a total candidate of 1456 discharge summaries.

**Table 1.** Selected phenotypes and their definitions.

| Phenotype | Definition |
| --- | --- |
| **High Prevalence** |  |
| Depression | Mention of depression in the patient’s medical history OR prescription of antidepressants. |
| Type II Diabetes | Mention of a history of type II diabetes AND on oral medication OR insulin therapy. |
| Hypertension (essential) | Mention of a history of hypertension AND on antihypertensive medication. |
| **Low Prevalence** |  |
| Dementia | Mention of dementia due to Alzheimer’s disease, alcohol, or any other form of dementia. |
| Type I Diabetes | Mention of a history of type I diabetes AND on insulin therapy. |
| Heart Failure (preserved ejection fraction) | Mention of a history of congestive heart failure AND ejection fraction $>40\%$. |
| Heart Failure (reduced ejection fraction) | Mention of a history of congestive heart failure AND ejection fraction $\leq 40\%$. |
| Metastatic Cancer | Mention of cancer with very high/imminent mortality (e.g., cholangiocarcinoma). |
| Rheumatoid Arthritis | Mention of rheumatoid arthritis in the patient’s medical history AND description of the disease. |
| Systemic Lupus | Mention of lupus in the patient’s medical history. |
| **Lifestyle-Associated Conditions** |  |
| Alcohol Abuse | Description of $>7$ alcoholic drinks per week OR complications due to alcohol (alcoholic hepatitis, alcohol intoxication, alcohol withdrawal). |
| Obesity | Body mass index (BMI) $>30$ OR being considered for/undergoing bariatric surgery. |
| **Healthcare-Associated Events** |  |
| C. difficile (complication) | Mention of C. difficile during the hospital stay. |
| C. difficile (medical history) | Mention of C. difficile in the patient’s medical history. |
| Deep Vein Thrombosis (DVT) / Pulmonary Embolism (PE) (complication) | Mention of DVT or PE occurring during the hospital stay. |
| DVT/PE (medical history) | Mention of DVT or PE in the patient’s medical history. |

Two annotators, one internal medicine physician and one medical student independently reviewed each candidate 1456 discharge summaries using the annotation software Labelbox^19^. This was a multilabel annotation task as each discharge summary was annotated for the presence or absence of each of the sixteen phenotypes. We provided annotators with a set of comprehensive a priori definitions for each phenotype (see Table 1). Where explicit mention of a diagnosis was absent, we allowed for indirect evidence to guide annotation.

Following the independent pass, both annotators jointly reviewed every case with any disagreement, engaging in open deliberation to resolve conflict. In instances where neither annotator found sufficient evidence to support a definitive label after discussion, the entire document was rejected. We did not systematically document the rationale for every decision or record detailed comment logs during consensus rounds.

We assessed inter-annotator agreement between annotators using Cohen’s Kappa ()^20^. In the context of multi-label annotations, we computed agreement at two levels: per-document (agreement across all labels within each discharge summary) and per-label (agreement for each phenotype across all documents)^21^. Document-level reflects holistic agreement on the entire multi-class classification per discharge summary, while label-level reflects how agreement varies for a given phenotype. We present the averaged scores across all discharge summaries and across all labels respectively. To contextualize disagreements at the document level, we also recorded the total number of conflicting labels per document and categorized disagreements into two types: (1) labels assigned by the physician but omitted by the student (physician-only positives) and (2) labels assigned by the student but omitted by the physician (student-only positives). We also calculated label disagreement rates as the proportion of conflicting annotations relative to the total annotated labels (union of positive labels by the two annotators).

To determine which discharge summaries were included, we followed the inclusion rule presented in Figure 2. Discharge summaries with perfect consensus (identical classification for all labels) were directly accepted. Those with three or more disagreements were rejected. Documents with one or two disagreements were reviewed conjointly by the two annotators for a final consensus review. If no consensus could be reached, the discharge summary was rejected, otherwise the document was accepted with the revised labels.

**Figure 2.**
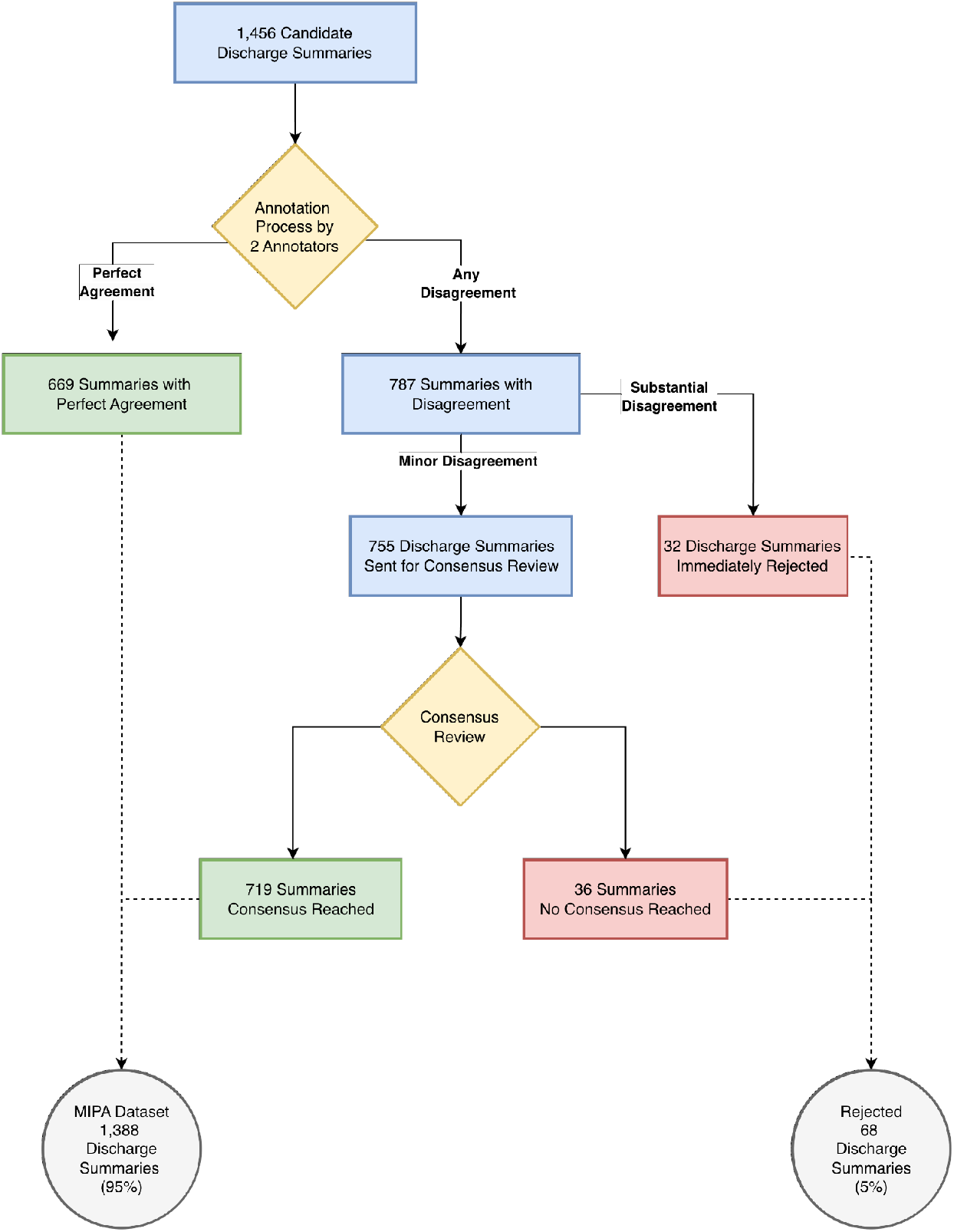
Flow Diagram of the MIPA Dataset Creation. *Note*. Flowchart depicting the process of creating the MIPA dataset. Initial review of 1,456 discharge summaries led to 669 summaries with perfect annotation agreement and 755 requiring consensus review. A disagreement threshold of three or more discordant labels per discharge summary resulted in rejection.

**Figure 3.**
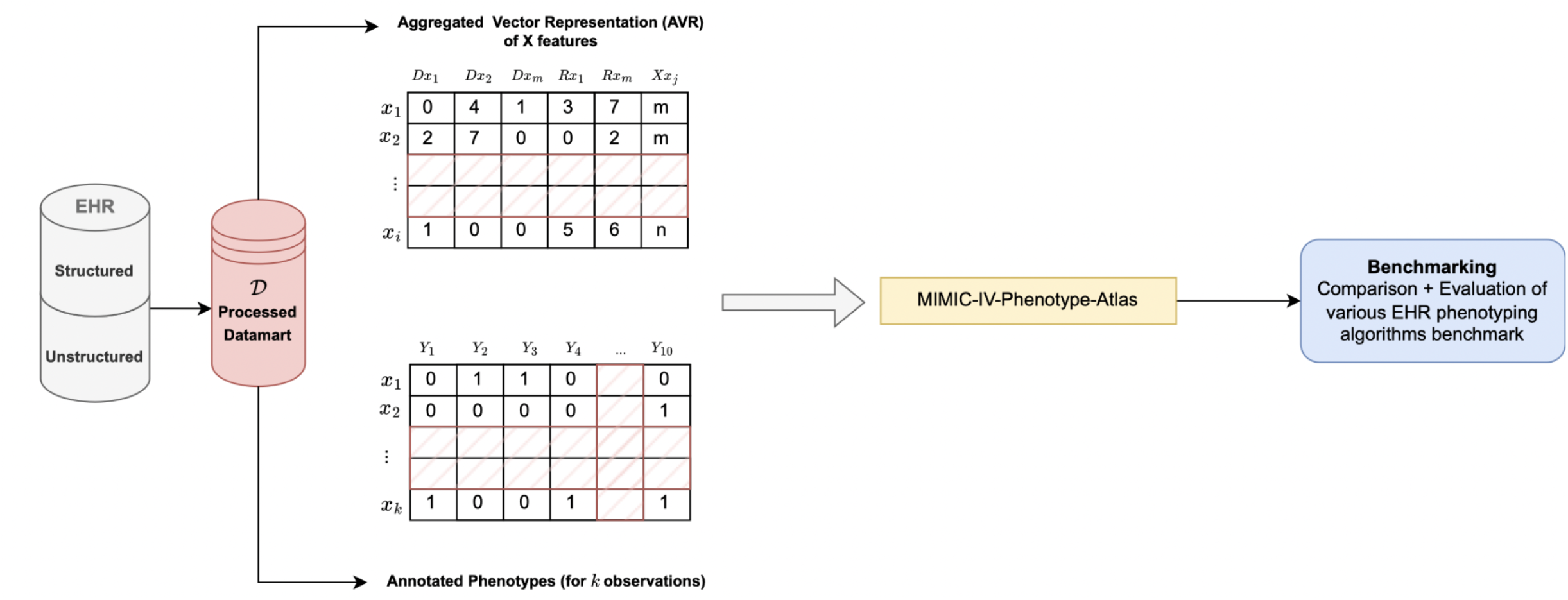
Figure showing the applicability and usability of the MIPA phenotyping database *Note*. MIMIC-IV in its raw form is a SQL database of 44 distinct tables (details available at https://mimic.mit.edu/docs/iv/). For the specific task of EHR phenotyping, we selected, pre-processed, and aggregated the data relevant to the task, with the objective of transforming MIMIC-IV into a patient-level vectorized format suitable for prediction tasks.

### 1.2.3 Supervised Training Dataset

To prepare the MIMIC-IV dataset for supervised EHR phenotyping, we built a processing pipeline, adapted from the framework described in Gupta et al^22^. This pipeline consists of two major phases: cohort construction and data preprocessing.

#### 1) Cohort Construction

To leverage the full scale of the MIMIC-IV database beyond our manually annotated dataset of 1,456 records, we implemented a hybrid silver/gold labeling strategy. Because exhaustive manual chart review of all MIMIC-IV discharge summaries is prohibitively resource-intensive, we utilized weak supervised learning—a method previously validated for computational phenotyping^23,6^.

For the training set, we generated silver labels across the broader, unannotated MIMIC-IV database using a rule-based proxy. An admission was labeled positive for a given phenotype if it contained at least one corresponding ICD code (Supplementary File 1). Conversely, control (silver-negative) cases were drawn from admissions strictly lacking any ICD codes related to the target phenotype. To reduce class imbalance, we balanced the training set 50/50.

We exclusively reserved the 1,456 manually annotated discharge summaries (gold labels) for the validation and testing tests. Concretely, 40% for validation and 60% for testing. We did not artificially balance classes, respecting the original distribution of phenotypes in the dataset.

#### 2) Data Preprocessing and Feature Engineering

Data preprocessing aimed to harmonize raw EHR data into structured features suitable for supervised learning. This included feature reduction, entity extraction and across diagnoses, medications, labs, chart events, and clinical text.

All ICD-9 codes were mapped to ICD-10 by using the first three digits of ICD-10 codes as the unifying root. Medication names were normalized by mapping National Drug Codes (NDCs) identifiers to their generic names. Both steps allowed to reduce dimension without sacrificing clinical granularity.

We applied the MedCAT parser to all available clinical notes (discharge summaries and radiology reports) to extract UMLS Concept Unique Identifiers (CUIs)^24,25^. For each observation, extracted CUIs were aggregated into frequency counts, yielding high-dimensional structured text features.

To further control dimensionality and reduce noise, all features with a frequency below the 25th percentile within each phenotype-specific cohort. This includes procedure and diagnostic ICD codes, medications, chart events (those are routine bedside clinical observations such as vital signs, neurological status, ventilator settings, and laboratory events). For UMLS CUI concepts, the threshold was lower and all features below the 10th percentile frequency were removed. Similarly, percentile thresholds were computed within each phenotypic cohort.

For high-frequency tables (medications, labs, and chart events), the pipeline allows two options: aggregate counts of occurrences per admission or time-series aggregation, where events are summarized into hourly, daily, or weekly windows.

The final output is a pivoted, admission-level feature table for each phenotype, as each has its own positive and control observations (silver labels for training; gold labels for validation/testing).

### 1.2.4 Baseline EHR Phenotyping

We implemented two interpretable baseline methods. The first leveraged structured diagnostic data, using the curated list of ICD codes (see Supplementary File 1). For each observation, we calculated the count of matching ICD codes and generated binary classifications using three thresholds (≥1, ≥2, ≥3 codes). The second baseline method directly leveraged the discharge summaries. We employed a keyword-driven term frequency–inverse document frequency (TF-IDF) classifier for each phenotype (see Supplementary File 2)^26^. First, a TF-IDF matrix was computed over using the validation set of gold-labeled discharge summaries. Second, for each phenotype, we developed a curated list of keywords related to the phenotype (synonyms, abbreviations, misspellings; Supplementary Table S1) and computed a document-level relevance score equal to the sum of TF-IDF weights for all features containing these keywords. Binary predictions were generated by applying percentile-based thresholds (25th, 50th, 75th, 90th) to the distribution of relevance scores for that phenotype. Predictions were evaluated against the gold labels using the same metrics as the other baselines.

### 1.2.5 Supervised Machine Learning

To illustrate MIPA’s ability to support structured ML approaches, we trained four supervised learning models: logistic regression (LR), naive bayes (NB), random forest (RF), and gradient boosting (GB). Models were trained on the silver-labeled training set, with gold-standard annotations used exclusively for validation and testing. Evaluation was performed on the held-out test set using standard classification metrics (macroF1, precision, recall, specificity). Technical details regarding additional preprocessing steps, imputation, and hyperparameter tuning procedures are provided in Supplementary File 3.

### 1.2.6 Large Language Models

We evaluated LLMs for EHR phenotyping using GPT-4o^27^. Phenotype prediction was obtained prompting the model to classify an observation for the presence or absence of a phenotype provided the raw discharge summary. We used chain-of-thought prompting^28^ and a fixed temperature = 0.5. Evaluation was performed on the test set labels using the same metrics as above. Details on the system and instruction prompts used were included in Supplementary File 3.

## 1.3 Results

### 1.3.1 MIPA Dataset Annotation

We reviewed a total of 1456 candidate discharge summaries, of which 1388 were retained after consensus review (see Figure 2). Inter-annotator agreement was strong overall, with a mean document-level of 0.805 (near-perfect agreement) and a mean label-level of 0.771 (substantial agreement) across 16 phenotypes (see Table S2). Inter-annotator disagreements totaled 1193 across 787 discharge summaries. 32 discharge summaries (2.8%) involved substantial disagreements (three or more discordant labels) and were thus not submitted to consensus review. The remaining 755 discharge summaries and 1154 conflicting labels underwent adjudication by consensus review, with ultimately 719 discharge summaries achieving consensus.

Label-level analysis revealed relative homogeneity in interobserver agreement across the different phenotypes (see Table S3). After the first annotation round, the average interobserver agreement across all phenotypes was moderately high (= 0.77± 0.18). Highest agreement was observed for conditions such as C. difficile (medical history) (= 0.921) and metastatic cancer (= 0.920). All phenotypes had a label-level > 0.70, with the two exceptions being the none label (= 0.505) and obesity (= 0.23). Disagreements were further resolved through iterative consensus discussions, with 96.7% (1154/1193) labels achieving agreement (see Table S4). Physicians’ judgments were adopted more frequently (760 vs. 394 medical student agreements).

### 1.3.2 MIPA Dataset Characteristics

The final MIPA dataset comprises 1,388 discharge summaries across sixteen distinct phenotypes (Table 2). Hypertension (essential) was the most prevalent condition, documented in 67.7% of summaries (n=940), followed by depression (48.9%, n=679) and Type II diabetes (28.3%, n=393). Heart-failure with reduced ejection fraction, systemic lupus and dementia were the three phenotypes with the lowest prevalence (respectively 8.8%, 8.1% and 7.3%). Incidence of healthcare-associated events was low (Deep Vein Thrombosis and Pulmonary Embolism [DVT/PE]): 2.8%, n=39; C. difficile: 3.0%, n=42). Only 2.5% of observations did not have any phenotype. Table 3 provides a detailed overview of the MIPA dataset, including structural characteristics of discharge summaries and key tabular EHR components—admission-level characteristics, available features, demographic distributions, and clinical outcomes.

**Table 2.** Phenotype distribution in the MIPA Dataset (n = 1,388 Discharge Summaries)

| Category | Phenotype | Total cases | Prevalence |
| --- | --- | --- | --- |
| **High Prevalence** |  |  |  |
|  | Hypertension (essential) | 940 | 67.70% |
|  | Depression | 679 | 48.90% |
|  | Type II Diabetes | 393 | 28.30% |
| **Low Prevalence** |  |  |  |
|  | Heart Failure (preserved ejection fraction) | 217 | 15.60% |
|  | Metastatic Cancer | 168 | 12.10% |
|  | Rheumatoid Arthritis | 164 | 11.80% |
|  | Heart Failure (reduced ejection fraction) | 122 | 8.80% |
|  | Systemic Lupus | 113 | 8.10% |
|  | Dementia | 102 | 7.30% |
| **Lifestyle-Associated Conditions** |  |  |  |
|  | Obesity | 286 | 20.60% |
|  | Alcohol Abuse | 155 | 11.20% |
| **Healthcare-Associated Events** |  |  |  |
|  | DVT/PE (past history) | 312 | 22.50% |
|  | DVT/PE (complication) | 39 | 2.80% |
|  | C. difficile (past history) | 101 | 7.30% |
|  | C. difficile (complication) | 42 | 3.00% |
| **Other** |  |  |  |
|  | None | 35 | 2.50% |

*Note.* Conditions categorized by clinical relevance and prevalence. Prevalence reflects the proportion of discharge summaries containing each phenotype.

**Table 3.**
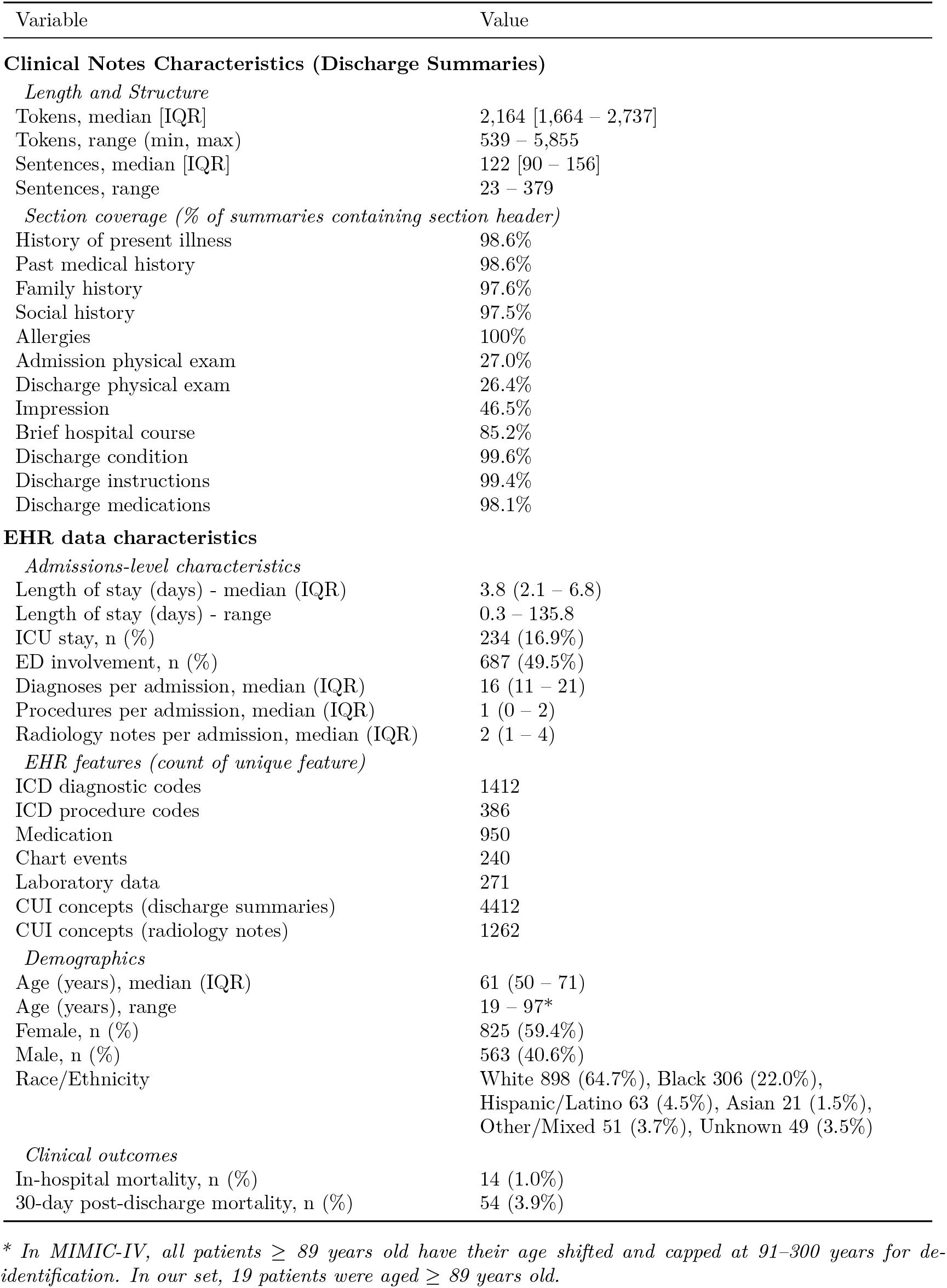
Characteristics of the MIMIC-IV Phenotype Atlas (MIPA) Dataset (n = 1,388 discharge summaries)

| Variable | Value |
| --- | --- |
| **Clinical Notes Characteristics (Discharge Summaries)** |  |
| *Length and Structure* |  |
| Tokens, median [IQR] | 2,164 [1,664 – 2,737] |
| Tokens, range (min, max) | 539 – 5,855 |
| Sentences, median [IQR] | 122 [90 – 156] |
| Sentences, range | 23 – 379 |
| *Section coverage (% of summaries containing section header)* |  |
| History of present illness | 98.6% |
| Past medical history | 98.6% |
| Family history | 97.6% |
| Social history | 97.5% |
| Allergies | 100% |
| Admission physical exam | 27.0% |
| Discharge physical exam | 26.4% |
| Impression | 46.5% |
| Brief hospital course | 85.2% |
| Discharge condition | 99.6% |
| Discharge instructions | 99.4% |
| Discharge medications | 98.1% |
| **EHR data characteristics** |  |
| *Admissions-level characteristics* |  |
| Length of stay (days) - median (IQR) | 3.8 (2.1 – 6.8) |
| Length of stay (days) - range | 0.3 – 135.8 |
| ICU stay, n (%) | 234 (16.9%) |
| ED involvement, n (%) | 687 (49.5%) |
| Diagnoses per admission, median (IQR) | 16 (11 – 21) |
| Procedures per admission, median (IQR) | 1 (0 – 2) |
| Radiology notes per admission, median (IQR) | 2 (1 – 4) |
| *EHR features (count of unique feature)* |  |
| ICD diagnostic codes | 1412 |
| ICD procedure codes | 386 |
| Medication | 950 |
| Chart events | 240 |
| Laboratory data | 271 |
| CUI concepts (discharge summaries) | 4412 |
| CUI concepts (radiology notes) | 1262 |
| *Demographics* |  |
| Age (years), median (IQR) | 61 (50 – 71) |
| Age (years), range | 19 – 97* |
| Female, n (%) | 825 (59.4%) |
| Male, n (%) | 563 (40.6%) |
| Race/Ethnicity | White 898 (64.7%), Black 306 (22.0%),<br>Hispanic/Latino 63 (4.5%), Asian 21 (1.5%),<br>Other/Mixed 51 (3.7%), Unknown 49 (3.5%) |
| *Clinical outcomes* |  |
| In-hospital mortality, n (%) | 14 (1.0%) |
| 30-day post-discharge mortality, n (%) | 54 (3.9%) |

*\* In MIMIC-IV, all patients $\geq 89$ years old have their age shifted and capped at 91–300 years for de-identification. In our set, 19 patients were aged $\geq 89$ years old.*

### 1.3.3 EHR Phenotyping Benchmarking Case Study

To illustrate MIPA’s utility as a benchmarking resource, we conducted a case study comparing ICD-based heuristics, keyword-driven TF-IDF classifiers, supervised machine learning models, and a large language model (GPT-4o) across the 16 phenotypes (Table 4, Table S5 for detailed results).

**Table 4.**
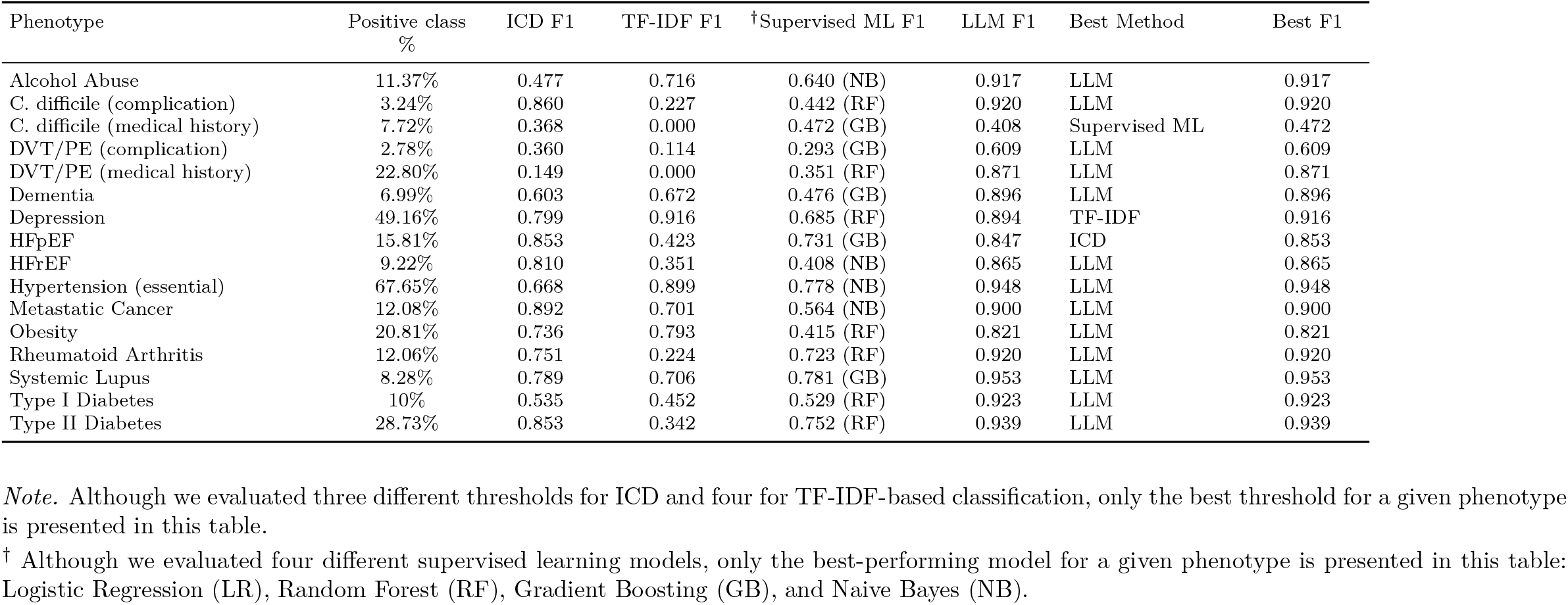
EHR Phenotyping Case Study Results.

Overall, model performance varied by phenotype prevalence and task complexity.

Baseline methods showed modest performance. ICD-based heuristics (F1 = 0.65± 0.23, for≥ threshold) performed well for conditions with reliable structured coding (e.g., HFpEF, HFrEF, metastatic cancer) but showed sharp performance declines at higher code thresholds (≥ 2, ≥ 3 with respective F1 being 0.13 and 0.07). Performance of keyword-driven TF-IDF classifiers was more heterogeneous across phenotypes (F1 = 0.43± 0.30 for the 25th percentile threshold). TF-IDF classifiers matched other methods for depression (F1 = 0.92), hypertension (F1 = 0.90), obesity (F1 = 0.78), alcohol abuse (F1 = 0.71) and metastatic cancer (F1 =0.70) but were mostly unreliable for the remainder phenotypes averaging an F1 0.26 ± 0.17).

Supervised ML did not show performance gains over baselines, with a F1= 0.44 ± 0.18 across all four learning methods. The best model differed by phenotype (e.g., GB for systemic lupus F1= 0.78, RF for type II diabetes F1=0.75, NB for hypertension F1= 0.78). Detailed results for all phenotypes are provided in Table S5.

LLM achieved the highest overall phenotyping performance (F1 = 0.85 ± 0.14). For 13 of 16 phenotypes, the LLM achieved the highest F1 score, with improvements over the next-best method ranging from +0.05 to +0.52. The largest margins were observed for DVT/PE medical history (+0.52), type 1 diabetes (+0.39), and systemic lupus (+0.16), all conditions where narrative descriptions contain nuanced contextual information not easily captured by tabular data or keywords. Importantly, performance gains were not explained by phenotype prevalence alone. While LLM performance gains were seen for low-prevalence phenotypes (e.g., Dementia at 3.2% prevalence, F1 = 0.90 an improvement from 0.68 with TF-IDF), substantial gains were also observed in moderate-prevalence conditions such as DVT/PE medical history (22.8%).

## 1.4 Discussion

### 1.4.1 MIPA Dataset

In this work, we introduce the MIMIC-IV Phenotype Atlas (MIPA), the first open-access, benchmark dataset specifically designed for electronic health record (EHR) phenotyping. By providing gold-standard expert-annotated labels for sixteen clinically relevant and diverse phenotypes, MIPA directly addresses a central barrier in the field: the absence of a standardized resource that enables reproducible evaluation and fair comparison of phenotyping methods.

Annotation quality is a foundational characteristic of any benchmark resource. In our annotation workflow (Figure 2), two independent annotators reviewed 1,456 candidate discharge summaries. Inter-annotator agreement was substantial before consensual review (document-level 0.805; label-level 0.755). Among the 787 discharge summaries with an initial discordance, consensual agreement was reached for 91%. A total of 71 summaries (5%) were excluded, 32 due to substantial disagreement and 36 that remained unresolved after consensual review, yielding a final corpus of 1,388 summaries. These numbers underscore both the strengths and the limitations of our annotation process. This high resolution rate and low rejection rate suggests that most phenotypes could be reliably identified from available documentation, while the unresolved cases highlight intrinsic ambiguities present in real-world clinical notes.

Pre-consensus agreement varied by phenotype (Table S3). Conditions with explicit diagnostic cues (e.g., C. difficile history, metastatic cancer) exhibited near-perfect agreement (0.92).

In contrast, phenotypes requiring inference (e.g. alcohol abuse, = 0.78), clinical judgment (e.g. obesity, = 0.22), or interpretation of test results (e.g. HFrEF, HFpEF = 0.69 and 0.79) showed lower agreement. Obesity exhibited the lowest pre-consensus agreement overwhelmingly driven by physician-only positives, which was due to different interpretation of diagnostic criteria by the two annotators: implicit inference from clinical description by the physician and reliance on explicit BMI documentation by the student. Importantly, this heterogeneity in label-level agreement, is not a limitation but rather a defining feature of a useful phenotyping benchmark: it captures the semantic ambiguity and variability that computational systems must navigate.

Complementing the annotated corpus, we provide a complete processing pipeline (publicly available on GitHub) that transforms raw MIMIC-IV data into phenotype-specific feature matrices suitable for benchmarking. The pipeline includes processed EHR data (diagnoses, procedures, medications, labs, chart events, and UMLS CUI extracted from clinical notes) and predefined training, validation and test set for the development of supervised learning based phenotyping methods.

### 1.4.2 Benchmarking Study

Our benchmarking case study illustrates how MIPA can be used to systematically evaluate diverse phenotyping strategies. While this analysis was designed to demonstrate benchmarking enabled by MIPA, rather than establish a leaderboard of methods, several patterns emerged.

First, baseline methods remained competitive for well-coded conditions such as heart failure, while keyword-driven TF-IDF approaches performed best for conditions with clear clinical documentation requiring minimal contextual reasoning such as hypertension and depression. Traditional supervised models yielded lower F1 scores than those reported in prior work. This difference reflects the intentionally conservative experimental setup of MIPA, which restricts feature extraction to the index admission rather than leveraging longitudinal aggregation or extensive feature engineering. In this context, reduced supervised performance highlights the intrinsic difficulty of EHR phenotyping when relying solely on contemporaneous structured data and underscores the importance of contextual and temporal information.

Large language models achieved the highest F1 scores across most phenotypes, suggesting that their ability to integrate unstructured narrative context enables performance gains precisely where traditional approaches are most constrained. Robustness to class imbalance alone did not explain LLM performance gains. Instead, observed improvements were shaped by two characteristics: weak or inconsistent structured EHR proxies and phenotypes whose evidence is primarily embedded within contextual and temporal narrative descriptions.

This explains why LLMs showed large gains for DVT/PE history, rheumatoid arthritis, and type I diabetes—even when prevalence was moderate—and smaller gains for phenotypes with strong structured signals such as metastatic cancer or hypertension, two phenotypes for which supervised learning methods performed well. This finding is consistent with previous EHR phenotyping work showing that the relative importance of text compared to tabular data depends on the phenotype in question with social determinants of health and behavioral phenotypes relying heavily on narrative documentation whereas biomarker-driven and relatively well coded conditions (for example, diabetes and hypertension) are often well captured by structured elements alone^29–31^.

### 1.4.3 Limitations

Several limitations warrant consideration. First, our annotation process while rigorous did not include independent third-party adjudication, and detailed decision logs were not preserved. Most disagreements stemmed from interpretive differences of inclusion criteria for obesity (e.g., qualitative descriptors of obesity) and instances where phenotypes were inferred (e.g., inferring HFrEF from medications or devices), or occasional omission by annotators. Differences in comfort with clinical inference between physician and student annotators contributed to these discrepancies.

Second, MIPA reflects data from a single academic health system, limiting generalizability across institutions and care settings. Phenotype prevalences also differ from real-world epidemiology due to sampling constraints and annotation scope. Finally, although discharge summaries offer rich narrative content, they represent only one component of longitudinal patient records; in the context of MIMIC-IV, this was the only clinical notes available.

## 1.5 Conclusion

In summary, MIPA establishes the first openly available benchmark dataset for EHR phenotyping, combining expert annotations, diverse phenotypes, and transparent case study evaluations. By enabling reproducible and objective comparison of methods, MIPA aims to catalyze progress in automated phenotyping and provide the community with a durable reference point for future research.

## Supporting information

Supplemental File 1 - Silver Labels

Supplemental File 2 - TF-IDF Keywords

Supplemental File 3- Supervised Learning Methods and Large Language Model Pipeline

Supplemental File 4 - ACCORD checklist table

## Data Availability

The MIMIC-IV Phenotype Atlas (MIPA) dataset labels are openly available at https://github.com/open-health-data-lab/MIPA-datacard

All code for pipeline processing, supervised model training, and LLM-based phenotyping is available at https://github.com/open-health-data-lab/MIPA

Note that access to the underlying discharge summaries and clinical notes is conditional upon completing CITI training and obtaining authorized access to the MIMIC-IV v2.2 database through PhysioNet.

## Abbreviation

CUI: (Concept Unique Identifier)
EHR: (Electronic Health Record)
ICD: (Inter-national Classification of Diseases)
LLM: (Large Language Model)
MIPA: (MIMIC-IV Phenotype Atlas)
ML: (Machine Learning)
TF-IDF: (Term Frequency–Inverse Document Frequency)
UMLS: (Unified Medical Language System),

## Acknowledgment

Portions of the analysis pipeline and this manuscript were drafted and edited with the assistance of a large language model.

## References

1. Banerji A, Blumenthal KG, Lai KH, Zhou L. Epidemiology of ACE inhibitor angioedema utilizing a large electronic health record. The Journal of Allergy and Clinical Immunology: In Practice 2017;5(3):744–9.

2. Denny JC, Ritchie MD, Basford MA, et al. PheWAS: demonstrating the feasibility of a phenome-wide scan to discover gene-disease associations. Bioinformatics 2010;26(9):1205–10.

3. Huang F, Hou J, Zhou N, et al. Advancing the Use of Longitudinal Electronic Health Records: Tutorial for Uncovering Real-World Evidence in Chronic Disease Outcomes. Journal of Medical Internet Research 2025;27:e71873.

4. Yu S, Ma Y, Gronsbell J, et al. Enabling phenotypic big data with PheNorm. Journal of the American Medical Informatics Association 2017;0(0):7.

5. Zhang Y, Cai T, Yu S, et al. High-throughput phenotyping with electronic medical record data using a common semi-supervised approach (PheCAP). Nature Protocols 2019;14(12):3426–44.

6. Halpern Y, Horng S, Choi Y, Sontag D. Electronic medical record phenotyping using the anchor and learn framework. Journal of the American Medical Informatics Association 2016;23(4):731–40.

7. Banda JM, Seneviratne M, Hernandez-Boussard T, Shah NH. Advances in electronic phenotyping: from rule-based definitions to machine learning models. Annual Review of Biomedical Data Science 2018;1(1):53–68.

8. Owens D, Nguyen DQ, Dohopolski M, Rousseau JF, Peterson ED, Navar AM. Accuracy of large language models to identify stroke subtypes within unstructured electronic health record data. Stroke 2025;56(10):2966–75.

9. Yan C, Ong HH, Grabowska ME, et al. Large language models facilitate the generation of electronic health record phenotyping algorithms. Journal of the American Medical Informatics Association 2024;31(9):1994–2001.

10. He T, Belouali A, Patricoski J, et al. Trends and opportunities in computable clinical phenotyping: A scoping review. Journal of Biomedical Informatics 2023;140:104335.

11. Uzuner O, Goldstein I, Luo Y, Kohane I. Identifying patient smoking status from medical discharge records. J Am Med Inform Assoc 2008;15(1):14–24.

12. Uzuner O. Recognizing obesity and comorbidities in sparse data. J Am Med Inform Assoc 2009;16(4):561–70.

13. Stubbs A, Filannino M, Soysal E, Henry S, Uzuner O. Cohort selection for clinical trials: n2c2 2018 shared task track 1. J Am Med Inform Assoc 2019;26(11):1163–71.

14. Wornow M, Thapa R, Steinberg E, Fries JA, Shah NH. EHRSHOT: An EHR Benchmark for Few-Shot Evaluation of Foundation Models [Internet]. 2023. Available from: https://arxiv.org/abs/2307.02028

15. Pollard TJ, Johnson AE, Raffa JD, Celi LA, Mark RG, Badawi O. The eICU Collaborative Research Database, a freely available multi-center database for critical care research. Scientific data 2018;5.

16. Johnson AEW, Bulgarelli L, Shen L. MIMIC-IV, a freely accessible electronic health record dataset. Sci Data 2023;10(1):1.

17. U.S. Department of Health and Human Services. Guidance Regarding Methods for Deidentification of Protected Health Information in Accordance with the HIPAA Privacy Rule [Internet]. Available from: https://www.hhs.gov/hipaa/for-professionals/privacy/special-topics/de-identification/index.html

18. Gattrell W. ACcurate COnsensus Reporting Document (ACCORD): a reporting guideline for consensus methods in biomedical research. PLoS Med 2024;21(1):e1004326.

19. Labelbox | The data factory for AI teams [Internet]. [cited 2025 Feb 23];Available from: https://labelbox.com

20. Hsu LM, Field R. Interrater agreement measures: Comments on Kappan, Cohen’s Kappa, Scott’s, and Aickin’s. Understanding Statistics 2003;2(3):205–19.

21. Ravenscroft J, Oellrich A, Saha S, Liakata M. Multi-label Annotation in Scientific Articles - The Multi-label Cancer Risk Assessment Corpus [Internet]. In: Calzolari N, Choukri K, Declerck T, et al., editors. Proceedings of the Tenth International Conference on Language Resources and Evaluation (LREC’16). Portorož, Slovenia: European Language Resources Association (ELRA); 2016 [cited 2026 Feb 4]. p. 4115–23.Available from: https://aclanthology.org/L16-1650/

22. Gupta M, Gallamoza B, Cutrona N, Dhakal P, Poulain R, Beheshti R. An Extensive Data Processing Pipeline for MIMIC-IV [Internet]. In: Proceedings of the 2nd Machine Learning for Health symposium. PMLR; 2022 [cited 2025 Sept 4]. p. 311–25. Available from: https://proceedings.mlr.press/v193/gupta22a.html

23. Nogues I-E, Wen J, Lin Y, et al. Weakly Semi-supervised phenotyping using Electronic Health records. Journal of Biomedical Informatics 2022;134:104175.

24. Kraljevic Z, Searle T, Shek A, et al. Multi-domain clinical natural language processing with MedCAT: The Medical Concept Annotation Toolkit. Artificial Intelligence in Medicine 2021;117:102083.

25. Rasmy L, Tiryaki F, Zhou Y, et al. Representation of EHR data for predictive modeling: a comparison between UMLS and other terminologies. J Am Med Inform Assoc 2020;27(10):1593–9.

26. Zeng Z, Deng Y, Li X, Naumann T, Luo Y. Natural language processing for EHR-based computational phenotyping. IEEE/ACM transactions on computational biology and bioinformatics 2018;16(1):139–53.

27. OpenAI. Hello GPT-4o [Internet]. 2024; Available from: https://openai.com/index/hello-gpt-4o/

28. Wei J, Wang X, Schuurmans D, et al. Chain-of-thought prompting elicits reasoning in large language models. Advances in neural information processing systems 2022;35:24824–37.

29. Moldwin E, Adekkanattu P, Rao S, et al. Empirical findings on the role of structured data, unstructured data, and their combination for automatic clinical phenotyping. AMIA Joint Summits on Translational Science proceedings 2021;2021:439–48.

30. Feller DJ, Zucker J, Yin MT, Gordon P, Elhadad N. Detecting social and behavioral determinants of health with structured and free-text clinical data. Applied Clinical Informatics 2020;11(1):1–8.

31. Wei W-Q, Teixeira PL, Mo H, Cronin RM, Warner JL, Denny JC. Combining billing codes, clinical notes, and medications from electronic health records provides superior phenotyping performance. Journal of the American Medical Informatics Association 2016;23(e1):e20–7.

