## Supplemental File 1 - Silver Labels for "MIMIC-IV Phenotype Atlas (MIPA): A Publicly Available Dataset for EHR Phenotyping"

### 1 Supplemental File 1 - Silver Labels

ICD Code Reference

Silver Label Phenotype Definitions

SA-1 Dataset · 16 Phenotypes

Phenotype Summary

| # | Phenotype | Total | ICD-9 | ICD-10 |
|---|---|---|---|---|
| 1 | C. difficile – Complication | 4 | 1 | 3 |
| 2 | C. difficile – Past History | 4 | 1 | 3 |
| 3 | Dementia | 23 | 14 | 9 |
| 4 | Depression | 60 | 38 | 22 |
| 5 | Diabetes Mellitus – Type 1 | 91 | 0 | 91 |
| 6 | Diabetes Mellitus – Type 2 | 453 | 66 | 387 |
| 7 | Heart Failure with Reduced Ejection Fraction (HFrEF) | 21 | 10 | 11 |
| 8 | Heart Failure with Preserved Ejection Fraction (HFpEF) | 9 | 5 | 4 |
| 9 | Hypertension | 8 | 3 | 5 |
| 10 | Systemic Lupus Erythematosus (SLE) | 18 | 7 | 11 |
| 11 | Metastatic Cancer | 86 | 28 | 58 |
| 12 | Obesity | 11 | 2 | 9 |
| 13 | Rheumatoid Arthritis | 422 | 8 | 414 |
| 14 | Alcohol Abuse / Use Disorder | 78 | 5 | 73 |
| 15 | Venous Thromboembolism (VTE) – Complication | 174 | 58 | 116 |
| 16 | Venous Thromboembolism (VTE) – Past History | 174 | 58 | 116 |

Detailed Code Listings

C. difficile – Complication

4 codes

| # | ICD Code | Version |
|---|---|---|
| 1 | A047 | ICD-10 |
| 2 | A0471 | ICD-10 |
| 3 | A0472 | ICD-10 |
| 4 | 00845 | ICD-9 |

C. difficile – Past History

4 codes

| # | ICD Code | Version |
|---|---|---|
| 1 | A047 | ICD-10 |
| 2 | A0471 | ICD-10 |
| 3 | A0472 | ICD-10 |
| 4 | 00845 | ICD-9 |

Dementia

23 codes

| # | ICD Code | Version |
|---|---|---|
| 1 | 2900 | ICD-9 |
| 2 | 29010 | ICD-9 |
| 3 | 29011 | ICD-9 |
| 4 | 29012 | ICD-9 |
| 5 | 29013 | ICD-9 |
| 6 | 29020 | ICD-9 |
| 7 | 29021 | ICD-9 |
| 8 | 2903 | ICD-9 |
| 9 | 29040 | ICD-9 |
| 10 | 29041 | ICD-9 |
| 11 | 29042 | ICD-9 |
| 12 | 29043 | ICD-9 |
| 13 | 2908 | ICD-9 |
| 14 | 2909 | ICD-9 |
| 15 | F00 | ICD-10 |
| 16 | F01 | ICD-10 |
| 17 | F02 | ICD-10 |
| 18 | F03 | ICD-10 |
| 19 | G30 | ICD-10 |
| 20 | G310 | ICD-10 |
| 21 | G3101 | ICD-10 |
| 22 | G3109 | ICD-10 |
| 23 | G311 | ICD-10 |

Depression

60 codes

| # | ICD Code | Version |
|---|---|---|
| 1 | 29620 | ICD-9 |
| 2 | 29621 | ICD-9 |
| 3 | 29622 | ICD-9 |
| 4 | 29623 | ICD-9 |
| 5 | 29624 | ICD-9 |
| 6 | 29625 | ICD-9 |
| 7 | 29626 | ICD-9 |
| 8 | 29630 | ICD-9 |

| # | ICD Code | Version |
|---|---|---|
| 9 | 29631 | ICD-9 |
| 10 | 29632 | ICD-9 |
| 11 | 29633 | ICD-9 |
| 12 | 29634 | ICD-9 |
| 13 | 29635 | ICD-9 |
| 14 | 29636 | ICD-9 |
| 15 | 29640 | ICD-9 |
| 16 | 29641 | ICD-9 |
| 17 | 29642 | ICD-9 |
| 18 | 29643 | ICD-9 |
| 19 | 29644 | ICD-9 |
| 20 | 29645 | ICD-9 |
| 21 | 29646 | ICD-9 |
| 22 | 29650 | ICD-9 |
| 23 | 29651 | ICD-9 |
| 24 | 29652 | ICD-9 |
| 25 | 29653 | ICD-9 |
| 26 | 29654 | ICD-9 |
| 27 | 29655 | ICD-9 |
| 28 | 29656 | ICD-9 |
| 29 | 2966 | ICD-9 |
| 30 | 29661 | ICD-9 |
| 31 | 29662 | ICD-9 |
| 32 | 29663 | ICD-9 |
| 33 | 29664 | ICD-9 |
| 34 | 29665 | ICD-9 |
| 35 | 29666 | ICD-9 |
| 36 | 3004 | ICD-9 |
| 37 | 3090 | ICD-9 |
| 38 | 311 | ICD-9 |
| 39 | F32 | ICD-10 |
| 40 | F320 | ICD-10 |
| 41 | F321 | ICD-10 |
| 42 | F322 | ICD-10 |
| 43 | F323 | ICD-10 |
| 44 | F328 | ICD-10 |
| 45 | F329 | ICD-10 |
| 46 | F33 | ICD-10 |
| 47 | F330 | ICD-10 |
| 48 | F331 | ICD-10 |
| 49 | F332 | ICD-10 |
| 50 | F333 | ICD-10 |
| 51 | F338 | ICD-10 |
| 52 | F339 | ICD-10 |
| 53 | F34 | ICD-10 |
| 54 | F340 | ICD-10 |
| 55 | F341 | ICD-10 |
| 56 | F348 | ICD-10 |
| 57 | F349 | ICD-10 |
| 58 | F38 | ICD-10 |
| 59 | F39 | ICD-10 |
| 60 | F41 | ICD-10 |

Diabetes Mellitus – Type 1

91 codes

| # | ICD Code | Version |
|---|---|---|
| 1 | E1010 | ICD-10 |
| 2 | E1011 | ICD-10 |
| 3 | E1021 | ICD-10 |
| 4 | E1022 | ICD-10 |
| 5 | E1029 | ICD-10 |
| 6 | E10311 | ICD-10 |
| 7 | E10319 | ICD-10 |
| 8 | E10321 | ICD-10 |
| 9 | E103211 | ICD-10 |
| 10 | E103212 | ICD-10 |
| 11 | E103213 | ICD-10 |
| 12 | E103219 | ICD-10 |
| 13 | E10329 | ICD-10 |
| 14 | E103291 | ICD-10 |
| 15 | E103292 | ICD-10 |
| 16 | E103293 | ICD-10 |
| 17 | E103299 | ICD-10 |
| 18 | E10331 | ICD-10 |
| 19 | E103311 | ICD-10 |
| 20 | E103312 | ICD-10 |
| 21 | E103313 | ICD-10 |
| 22 | E103319 | ICD-10 |
| 23 | E10339 | ICD-10 |
| 24 | E103391 | ICD-10 |
| 25 | E103392 | ICD-10 |
| 26 | E103393 | ICD-10 |
| 27 | E103399 | ICD-10 |
| 28 | E10341 | ICD-10 |
| 29 | E103411 | ICD-10 |
| 30 | E103412 | ICD-10 |
| 31 | E103413 | ICD-10 |
| 32 | E103419 | ICD-10 |
| 33 | E10349 | ICD-10 |
| 34 | E103491 | ICD-10 |
| 35 | E103492 | ICD-10 |
| 36 | E103493 | ICD-10 |
| 37 | E103499 | ICD-10 |
| 38 | E10351 | ICD-10 |
| 39 | E103511 | ICD-10 |
| 40 | E103512 | ICD-10 |
| 41 | E103513 | ICD-10 |
| 42 | E103519 | ICD-10 |
| 43 | E103521 | ICD-10 |
| 44 | E103522 | ICD-10 |
| 45 | E103523 | ICD-10 |
| 46 | E103529 | ICD-10 |
| 47 | E103531 | ICD-10 |
| 48 | E103532 | ICD-10 |
| 49 | E103533 | ICD-10 |

| # | ICD Code | Version |
|---|---|---|
| 50 | E103539 | ICD-10 |
| 51 | E103541 | ICD-10 |
| 52 | E103542 | ICD-10 |
| 53 | E103543 | ICD-10 |
| 54 | E103549 | ICD-10 |
| 55 | E103551 | ICD-10 |
| 56 | E103552 | ICD-10 |
| 57 | E103553 | ICD-10 |
| 58 | E103559 | ICD-10 |
| 59 | E10359 | ICD-10 |
| 60 | E103591 | ICD-10 |
| 61 | E103592 | ICD-10 |
| 62 | E103593 | ICD-10 |
| 63 | E103599 | ICD-10 |
| 64 | E1036 | ICD-10 |
| 65 | E1037X1 | ICD-10 |
| 66 | E1037X2 | ICD-10 |
| 67 | E1037X3 | ICD-10 |
| 68 | E1037X9 | ICD-10 |
| 69 | E1039 | ICD-10 |
| 70 | E1040 | ICD-10 |
| 71 | E1041 | ICD-10 |
| 72 | E1042 | ICD-10 |
| 73 | E1043 | ICD-10 |
| 74 | E1044 | ICD-10 |
| 75 | E1049 | ICD-10 |
| 76 | E1051 | ICD-10 |
| 77 | E1052 | ICD-10 |
| 78 | E1059 | ICD-10 |
| 79 | E10610 | ICD-10 |
| 80 | E10618 | ICD-10 |
| 81 | E10620 | ICD-10 |
| 82 | E10621 | ICD-10 |
| 83 | E10622 | ICD-10 |
| 84 | E10628 | ICD-10 |
| 85 | E10630 | ICD-10 |
| 86 | E10638 | ICD-10 |
| 87 | E10641 | ICD-10 |
| 88 | E10649 | ICD-10 |
| 89 | E1065 | ICD-10 |
| 90 | E1069 | ICD-10 |
| 91 | E108 | ICD-10 |

Diabetes Mellitus – Type 2

453 codes

| # | ICD Code | Version |
|---|---|---|
| 1 | E089 | ICD-10 |
| 2 | E099 | ICD-10 |
| 3 | E109 | ICD-10 |

| # | ICD Code | Version |
|---|---|---|
| 4 | E119 | ICD-10 |
| 5 | E139 | ICD-10 |
| 6 | R7301 | ICD-10 |
| 7 | R7302 | ICD-10 |
| 8 | R7303 | ICD-10 |
| 9 | R7309 | ICD-10 |
| 10 | R739 | ICD-10 |
| 11 | R81 | ICD-10 |
| 12 | R824 | ICD-10 |
| 13 | Z4681 | ICD-10 |
| 14 | Z9641 | ICD-10 |
| 15 | E0800 | ICD-10 |
| 16 | E0801 | ICD-10 |
| 17 | E0810 | ICD-10 |
| 18 | E0811 | ICD-10 |
| 19 | E0821 | ICD-10 |
| 20 | E0822 | ICD-10 |
| 21 | E0829 | ICD-10 |
| 22 | E08311 | ICD-10 |
| 23 | E08319 | ICD-10 |
| 24 | E08321 | ICD-10 |
| 25 | E083211 | ICD-10 |
| 26 | E083212 | ICD-10 |
| 27 | E083213 | ICD-10 |
| 28 | E083219 | ICD-10 |
| 29 | E08329 | ICD-10 |
| 30 | E083291 | ICD-10 |
| 31 | E083292 | ICD-10 |
| 32 | E083293 | ICD-10 |
| 33 | E083299 | ICD-10 |
| 34 | E08331 | ICD-10 |
| 35 | E083311 | ICD-10 |
| 36 | E083312 | ICD-10 |
| 37 | E083313 | ICD-10 |
| 38 | E083319 | ICD-10 |
| 39 | E08339 | ICD-10 |
| 40 | E083391 | ICD-10 |
| 41 | E083392 | ICD-10 |
| 42 | E083393 | ICD-10 |
| 43 | E083399 | ICD-10 |
| 44 | E08341 | ICD-10 |
| 45 | E083411 | ICD-10 |
| 46 | E083412 | ICD-10 |
| 47 | E083413 | ICD-10 |
| 48 | E083419 | ICD-10 |
| 49 | E08349 | ICD-10 |
| 50 | E083491 | ICD-10 |
| 51 | E083492 | ICD-10 |
| 52 | E083493 | ICD-10 |
| 53 | E083499 | ICD-10 |
| 54 | E08351 | ICD-10 |
| 55 | E083511 | ICD-10 |

| # | ICD Code | Version |
|---|---|---|
| 56 | E083512 | ICD-10 |
| 57 | E083513 | ICD-10 |
| 58 | E083519 | ICD-10 |
| 59 | E083521 | ICD-10 |
| 60 | E083522 | ICD-10 |
| 61 | E083523 | ICD-10 |
| 62 | E083529 | ICD-10 |
| 63 | E083531 | ICD-10 |
| 64 | E083532 | ICD-10 |
| 65 | E083533 | ICD-10 |
| 66 | E083539 | ICD-10 |
| 67 | E083541 | ICD-10 |
| 68 | E083542 | ICD-10 |
| 69 | E083543 | ICD-10 |
| 70 | E083549 | ICD-10 |
| 71 | E083551 | ICD-10 |
| 72 | E083552 | ICD-10 |
| 73 | E083553 | ICD-10 |
| 74 | E083559 | ICD-10 |
| 75 | E08359 | ICD-10 |
| 76 | E083591 | ICD-10 |
| 77 | E083592 | ICD-10 |
| 78 | E083593 | ICD-10 |
| 79 | E083599 | ICD-10 |
| 80 | E0836 | ICD-10 |
| 81 | E0837X1 | ICD-10 |
| 82 | E0837X2 | ICD-10 |
| 83 | E0837X3 | ICD-10 |
| 84 | E0837X9 | ICD-10 |
| 85 | E0839 | ICD-10 |
| 86 | E0840 | ICD-10 |
| 87 | E0841 | ICD-10 |
| 88 | E0842 | ICD-10 |
| 89 | E0843 | ICD-10 |
| 90 | E0844 | ICD-10 |
| 91 | E0849 | ICD-10 |
| 92 | E0851 | ICD-10 |
| 93 | E0852 | ICD-10 |
| 94 | E0859 | ICD-10 |
| 95 | E08610 | ICD-10 |
| 96 | E08618 | ICD-10 |
| 97 | E08620 | ICD-10 |
| 98 | E08621 | ICD-10 |
| 99 | E08622 | ICD-10 |
| 100 | E08628 | ICD-10 |
| 101 | E08630 | ICD-10 |
| 102 | E08638 | ICD-10 |
| 103 | E08641 | ICD-10 |
| 104 | E08649 | ICD-10 |
| 105 | E0865 | ICD-10 |
| 106 | E0869 | ICD-10 |
| 107 | E088 | ICD-10 |

| # | ICD Code | Version |
|---|---|---|
| 108 | E0900 | ICD-10 |
| 109 | E0901 | ICD-10 |
| 110 | E0910 | ICD-10 |
| 111 | E0911 | ICD-10 |
| 112 | E0921 | ICD-10 |
| 113 | E0922 | ICD-10 |
| 114 | E0929 | ICD-10 |
| 115 | E09311 | ICD-10 |
| 116 | E09319 | ICD-10 |
| 117 | E09321 | ICD-10 |
| 118 | E093211 | ICD-10 |
| 119 | E093212 | ICD-10 |
| 120 | E093213 | ICD-10 |
| 121 | E093219 | ICD-10 |
| 122 | E09329 | ICD-10 |
| 123 | E093291 | ICD-10 |
| 124 | E093292 | ICD-10 |
| 125 | E093293 | ICD-10 |
| 126 | E093299 | ICD-10 |
| 127 | E09331 | ICD-10 |
| 128 | E093311 | ICD-10 |
| 129 | E093312 | ICD-10 |
| 130 | E093313 | ICD-10 |
| 131 | E093319 | ICD-10 |
| 132 | E09339 | ICD-10 |
| 133 | E093391 | ICD-10 |
| 134 | E093392 | ICD-10 |
| 135 | E093393 | ICD-10 |
| 136 | E093399 | ICD-10 |
| 137 | E09341 | ICD-10 |
| 138 | E093411 | ICD-10 |
| 139 | E093412 | ICD-10 |
| 140 | E093413 | ICD-10 |
| 141 | E093419 | ICD-10 |
| 142 | E09349 | ICD-10 |
| 143 | E093491 | ICD-10 |
| 144 | E093492 | ICD-10 |
| 145 | E093493 | ICD-10 |
| 146 | E093499 | ICD-10 |
| 147 | E09351 | ICD-10 |
| 148 | E093511 | ICD-10 |
| 149 | E093512 | ICD-10 |
| 150 | E093513 | ICD-10 |
| 151 | E093519 | ICD-10 |
| 152 | E093521 | ICD-10 |
| 153 | E093522 | ICD-10 |
| 154 | E093523 | ICD-10 |
| 155 | E093529 | ICD-10 |
| 156 | E093531 | ICD-10 |
| 157 | E093532 | ICD-10 |
| 158 | E093533 | ICD-10 |
| 159 | E093539 | ICD-10 |

| # | ICD Code | Version |
|---|---|---|
| 160 | E093541 | ICD-10 |
| 161 | E093542 | ICD-10 |
| 162 | E093543 | ICD-10 |
| 163 | E093549 | ICD-10 |
| 164 | E093551 | ICD-10 |
| 165 | E093552 | ICD-10 |
| 166 | E093553 | ICD-10 |
| 167 | E093559 | ICD-10 |
| 168 | E09359 | ICD-10 |
| 169 | E093591 | ICD-10 |
| 170 | E093592 | ICD-10 |
| 171 | E093593 | ICD-10 |
| 172 | E093599 | ICD-10 |
| 173 | E0936 | ICD-10 |
| 174 | E0937X1 | ICD-10 |
| 175 | E0937X2 | ICD-10 |
| 176 | E0937X3 | ICD-10 |
| 177 | E0937X9 | ICD-10 |
| 178 | E0939 | ICD-10 |
| 179 | E0940 | ICD-10 |
| 180 | E0941 | ICD-10 |
| 181 | E0942 | ICD-10 |
| 182 | E0943 | ICD-10 |
| 183 | E0944 | ICD-10 |
| 184 | E0949 | ICD-10 |
| 185 | E0951 | ICD-10 |
| 186 | E0952 | ICD-10 |
| 187 | E0959 | ICD-10 |
| 188 | E09610 | ICD-10 |
| 189 | E09618 | ICD-10 |
| 190 | E09620 | ICD-10 |
| 191 | E09621 | ICD-10 |
| 192 | E09622 | ICD-10 |
| 193 | E09628 | ICD-10 |
| 194 | E09630 | ICD-10 |
| 195 | E09638 | ICD-10 |
| 196 | E09641 | ICD-10 |
| 197 | E09649 | ICD-10 |
| 198 | E0965 | ICD-10 |
| 199 | E0969 | ICD-10 |
| 200 | E098 | ICD-10 |
| 201 | E1010 | ICD-10 |
| 202 | E1111 | ICD-10 |
| 203 | E1121 | ICD-10 |
| 204 | E1122 | ICD-10 |
| 205 | E1129 | ICD-10 |
| 206 | E11311 | ICD-10 |
| 207 | E11319 | ICD-10 |
| 208 | E11321 | ICD-10 |
| 209 | E113211 | ICD-10 |
| 210 | E113212 | ICD-10 |
| 211 | E113213 | ICD-10 |

| # | ICD Code | Version |
|---|---|---|
| 212 | E113219 | ICD-10 |
| 213 | E11329 | ICD-10 |
| 214 | E113291 | ICD-10 |
| 215 | E113292 | ICD-10 |
| 216 | E113293 | ICD-10 |
| 217 | E113299 | ICD-10 |
| 218 | E11331 | ICD-10 |
| 219 | E113311 | ICD-10 |
| 220 | E113312 | ICD-10 |
| 221 | E113313 | ICD-10 |
| 222 | E113319 | ICD-10 |
| 223 | E11339 | ICD-10 |
| 224 | E113391 | ICD-10 |
| 225 | E113392 | ICD-10 |
| 226 | E113393 | ICD-10 |
| 227 | E113399 | ICD-10 |
| 228 | E11341 | ICD-10 |
| 229 | E113411 | ICD-10 |
| 230 | E113412 | ICD-10 |
| 231 | E113413 | ICD-10 |
| 232 | E113419 | ICD-10 |
| 233 | E11349 | ICD-10 |
| 234 | E113491 | ICD-10 |
| 235 | E113492 | ICD-10 |
| 236 | E113493 | ICD-10 |
| 237 | E113499 | ICD-10 |
| 238 | E11351 | ICD-10 |
| 239 | E113511 | ICD-10 |
| 240 | E113512 | ICD-10 |
| 241 | E113513 | ICD-10 |
| 242 | E113519 | ICD-10 |
| 243 | E113521 | ICD-10 |
| 244 | E113522 | ICD-10 |
| 245 | E113523 | ICD-10 |
| 246 | E113529 | ICD-10 |
| 247 | E113531 | ICD-10 |
| 248 | E113532 | ICD-10 |
| 249 | E113533 | ICD-10 |
| 250 | E113539 | ICD-10 |
| 251 | E113541 | ICD-10 |
| 252 | E113542 | ICD-10 |
| 253 | E113543 | ICD-10 |
| 254 | E113549 | ICD-10 |
| 255 | E113551 | ICD-10 |
| 256 | E113552 | ICD-10 |
| 257 | E113553 | ICD-10 |
| 258 | E113559 | ICD-10 |
| 259 | E11359 | ICD-10 |
| 260 | E113591 | ICD-10 |
| 261 | E113592 | ICD-10 |
| 262 | E113593 | ICD-10 |
| 263 | E113599 | ICD-10 |

| # | ICD Code | Version |
|---|---|---|
| 264 | E1136 | ICD-10 |
| 265 | E1137X1 | ICD-10 |
| 266 | E1137X2 | ICD-10 |
| 267 | E1137X3 | ICD-10 |
| 268 | E1137X9 | ICD-10 |
| 269 | E1139 | ICD-10 |
| 270 | E1140 | ICD-10 |
| 271 | E1141 | ICD-10 |
| 272 | E1142 | ICD-10 |
| 273 | E1143 | ICD-10 |
| 274 | E1144 | ICD-10 |
| 275 | E1149 | ICD-10 |
| 276 | E1151 | ICD-10 |
| 277 | E1152 | ICD-10 |
| 278 | E1159 | ICD-10 |
| 279 | E11610 | ICD-10 |
| 280 | E11618 | ICD-10 |
| 281 | E11620 | ICD-10 |
| 282 | E11621 | ICD-10 |
| 283 | E11622 | ICD-10 |
| 284 | E11628 | ICD-10 |
| 285 | E11630 | ICD-10 |
| 286 | E11638 | ICD-10 |
| 287 | E11641 | ICD-10 |
| 288 | E11649 | ICD-10 |
| 289 | E1165 | ICD-10 |
| 290 | E1169 | ICD-10 |
| 291 | E118 | ICD-10 |
| 292 | E1300 | ICD-10 |
| 293 | E1301 | ICD-10 |
| 294 | E1310 | ICD-10 |
| 295 | E1311 | ICD-10 |
| 296 | E1321 | ICD-10 |
| 297 | E1322 | ICD-10 |
| 298 | E1329 | ICD-10 |
| 299 | E13311 | ICD-10 |
| 300 | E13319 | ICD-10 |
| 301 | E13321 | ICD-10 |
| 302 | E133211 | ICD-10 |
| 303 | E133212 | ICD-10 |
| 304 | E133213 | ICD-10 |
| 305 | E133219 | ICD-10 |
| 306 | E13329 | ICD-10 |
| 307 | E133291 | ICD-10 |
| 308 | E133292 | ICD-10 |
| 309 | E133293 | ICD-10 |
| 310 | E133299 | ICD-10 |
| 311 | E13331 | ICD-10 |
| 312 | E133311 | ICD-10 |
| 313 | E133312 | ICD-10 |
| 314 | E133313 | ICD-10 |
| 315 | E133319 | ICD-10 |

| # | ICD Code | Version |
|---|---|---|
| 316 | E13339 | ICD-10 |
| 317 | E133391 | ICD-10 |
| 318 | E133392 | ICD-10 |
| 319 | E133393 | ICD-10 |
| 320 | E133399 | ICD-10 |
| 321 | E13341 | ICD-10 |
| 322 | E133411 | ICD-10 |
| 323 | E133412 | ICD-10 |
| 324 | E133413 | ICD-10 |
| 325 | E133419 | ICD-10 |
| 326 | E13349 | ICD-10 |
| 327 | E133491 | ICD-10 |
| 328 | E133492 | ICD-10 |
| 329 | E133493 | ICD-10 |
| 330 | E133499 | ICD-10 |
| 331 | E13351 | ICD-10 |
| 332 | E133511 | ICD-10 |
| 333 | E133512 | ICD-10 |
| 334 | E133513 | ICD-10 |
| 335 | E133519 | ICD-10 |
| 336 | E133521 | ICD-10 |
| 337 | E133522 | ICD-10 |
| 338 | E133523 | ICD-10 |
| 339 | E133529 | ICD-10 |
| 340 | E133531 | ICD-10 |
| 341 | E133532 | ICD-10 |
| 342 | E133533 | ICD-10 |
| 343 | E133539 | ICD-10 |
| 344 | E133541 | ICD-10 |
| 345 | E133542 | ICD-10 |
| 346 | E133543 | ICD-10 |
| 347 | E133549 | ICD-10 |
| 348 | E133551 | ICD-10 |
| 349 | E133552 | ICD-10 |
| 350 | E133553 | ICD-10 |
| 351 | E133559 | ICD-10 |
| 352 | E13359 | ICD-10 |
| 353 | E133591 | ICD-10 |
| 354 | E133592 | ICD-10 |
| 355 | E133593 | ICD-10 |
| 356 | E133599 | ICD-10 |
| 357 | E1336 | ICD-10 |
| 358 | E1337X1 | ICD-10 |
| 359 | E1337X2 | ICD-10 |
| 360 | E1337X3 | ICD-10 |
| 361 | E1337X9 | ICD-10 |
| 362 | E1339 | ICD-10 |
| 363 | E1340 | ICD-10 |
| 364 | E1341 | ICD-10 |
| 365 | E1342 | ICD-10 |
| 366 | E1343 | ICD-10 |
| 367 | E1344 | ICD-10 |

| # | ICD Code | Version |
|---|---|---|
| 368 | E1349 | ICD-10 |
| 369 | E1351 | ICD-10 |
| 370 | E1352 | ICD-10 |
| 371 | E1359 | ICD-10 |
| 372 | E13610 | ICD-10 |
| 373 | E13618 | ICD-10 |
| 374 | E13620 | ICD-10 |
| 375 | E13621 | ICD-10 |
| 376 | E13622 | ICD-10 |
| 377 | E13628 | ICD-10 |
| 378 | E13630 | ICD-10 |
| 379 | E13638 | ICD-10 |
| 380 | E13641 | ICD-10 |
| 381 | E13649 | ICD-10 |
| 382 | E1365 | ICD-10 |
| 383 | E1369 | ICD-10 |
| 384 | E138 | ICD-10 |
| 385 | 24900 | ICD-9 |
| 386 | 25000 | ICD-9 |
| 387 | 25001 | ICD-9 |
| 388 | 7902 | ICD-9 |
| 389 | 79021 | ICD-9 |
| 390 | 79022 | ICD-9 |
| 391 | 79029 | ICD-9 |
| 392 | 7915 | ICD-9 |
| 393 | 7916 | ICD-9 |
| 394 | V4585 | ICD-10 |
| 395 | V5391 | ICD-10 |
| 396 | V6546 | ICD-10 |
| 397 | 24901 | ICD-9 |
| 398 | 24910 | ICD-9 |
| 399 | 24911 | ICD-9 |
| 400 | 25002 | ICD-9 |
| 401 | 25003 | ICD-9 |
| 402 | 25010 | ICD-9 |
| 403 | 25011 | ICD-9 |
| 404 | 25012 | ICD-9 |
| 405 | 25013 | ICD-9 |
| 406 | 24940 | ICD-9 |
| 407 | 24941 | ICD-9 |
| 408 | 25040 | ICD-9 |
| 409 | 25041 | ICD-9 |
| 410 | 25042 | ICD-9 |
| 411 | 25043 | ICD-9 |
| 412 | 24950 | ICD-9 |
| 413 | 24951 | ICD-9 |
| 414 | 25050 | ICD-9 |
| 415 | 25051 | ICD-9 |
| 416 | 25052 | ICD-9 |
| 417 | 25053 | ICD-9 |
| 418 | 24960 | ICD-9 |
| 419 | 24961 | ICD-9 |

| # | ICD Code | Version |
|---|---|---|
| 420 | 25060 | ICD-9 |
| 421 | 25061 | ICD-9 |
| 422 | 25062 | ICD-9 |
| 423 | 25063 | ICD-9 |
| 424 | 24970 | ICD-9 |
| 425 | 24971 | ICD-9 |
| 426 | 25070 | ICD-9 |
| 427 | 25071 | ICD-9 |
| 428 | 25072 | ICD-9 |
| 429 | 25073 | ICD-9 |
| 430 | 24990 | ICD-9 |
| 431 | 24991 | ICD-9 |
| 432 | 25090 | ICD-9 |
| 433 | 25091 | ICD-9 |
| 434 | 24920 | ICD-9 |
| 435 | 24921 | ICD-9 |
| 436 | 24930 | ICD-9 |
| 437 | 24931 | ICD-9 |
| 438 | 24980 | ICD-9 |
| 439 | 24981 | ICD-9 |
| 440 | 25020 | ICD-9 |
| 441 | 25021 | ICD-9 |
| 442 | 25022 | ICD-9 |
| 443 | 25023 | ICD-9 |
| 444 | 25030 | ICD-9 |
| 445 | 25031 | ICD-9 |
| 446 | 25032 | ICD-9 |
| 447 | 25033 | ICD-9 |
| 448 | 25080 | ICD-9 |
| 449 | 25081 | ICD-9 |
| 450 | 25082 | ICD-9 |
| 451 | 25083 | ICD-9 |
| 452 | 25092 | ICD-9 |
| 453 | 25093 | ICD-9 |

Heart Failure with Reduced Ejection Fraction (HFrEF)

21 codes

| # | ICD Code | Version |
|---|---|---|
| 1 | 4282 | ICD-9 |
| 2 | 42820 | ICD-9 |
| 3 | 42821 | ICD-9 |
| 4 | 42822 | ICD-9 |
| 5 | 42823 | ICD-9 |
| 6 | 4284 | ICD-9 |
| 7 | 42840 | ICD-9 |
| 8 | 42841 | ICD-9 |
| 9 | 42842 | ICD-9 |
| 10 | 42843 | ICD-9 |
| 11 | I501 | ICD-10 |

| # | ICD Code | Version |
|---|---|---|
| 12 | I5020 | ICD-10 |
| 13 | I5021 | ICD-10 |
| 14 | I5022 | ICD-10 |
| 15 | I5023 | ICD-10 |
| 16 | I504 | ICD-10 |
| 17 | I5040 | ICD-10 |
| 18 | I5041 | ICD-10 |
| 19 | I5042 | ICD-10 |
| 20 | I5043 | ICD-10 |
| 21 | I509 | ICD-10 |

Heart Failure with Preserved Ejection Fraction (HFpEF)

9 codes

| # | ICD Code | Version |
|---|---|---|
| 1 | 4283 | ICD-9 |
| 2 | 42830 | ICD-9 |
| 3 | 42831 | ICD-9 |
| 4 | 42832 | ICD-9 |
| 5 | 42833 | ICD-9 |
| 6 | I503 | ICD-10 |
| 7 | I5031 | ICD-10 |
| 8 | I5032 | ICD-10 |
| 9 | I5033 | ICD-10 |

Hypertension

8 codes

| # | ICD Code | Version |
|---|---|---|
| 1 | 4010 | ICD-9 |
| 2 | 4011 | ICD-9 |
| 3 | 4019 | ICD-9 |
| 4 | I10 | ICD-10 |
| 5 | I11 | ICD-10 |
| 6 | I12 | ICD-10 |
| 7 | I13 | ICD-10 |
| 8 | I15 | ICD-10 |

Systemic Lupus Erythematosus (SLE)

18 codes

| # | ICD Code | Version |
|---|---|---|
| 1 | 7100 | ICD-9 |
| 2 | 7101 | ICD-9 |
| 3 | 7102 | ICD-9 |
| 4 | 7103 | ICD-9 |

| # | ICD Code | Version |
|---|---|---|
| 5 | 7104 | ICD-9 |
| 6 | 7108 | ICD-9 |
| 7 | 7109 | ICD-9 |
| 8 | M32 | ICD-10 |
| 9 | M320 | ICD-10 |
| 10 | M3210 | ICD-10 |
| 11 | M3211 | ICD-10 |
| 12 | M3212 | ICD-10 |
| 13 | M3213 | ICD-10 |
| 14 | M3214 | ICD-10 |
| 15 | M3215 | ICD-10 |
| 16 | M3219 | ICD-10 |
| 17 | M328 | ICD-10 |
| 18 | M329 | ICD-10 |

Metastatic Cancer

86 codes

| # | ICD Code | Version |
|---|---|---|
| 1 | 1960 | ICD-9 |
| 2 | 1961 | ICD-9 |
| 3 | 1962 | ICD-9 |
| 4 | 1963 | ICD-9 |
| 5 | 1965 | ICD-9 |
| 6 | 1966 | ICD-9 |
| 7 | 1968 | ICD-9 |
| 8 | 1970 | ICD-9 |
| 9 | 1971 | ICD-9 |
| 10 | 1972 | ICD-9 |
| 11 | 1973 | ICD-9 |
| 12 | 1974 | ICD-9 |
| 13 | 1975 | ICD-9 |
| 14 | 1976 | ICD-9 |
| 15 | 1977 | ICD-9 |
| 16 | 1978 | ICD-9 |
| 17 | 1983 | ICD-9 |
| 18 | 1984 | ICD-9 |
| 19 | 1985 | ICD-9 |
| 20 | 1986 | ICD-9 |
| 21 | 1987 | ICD-9 |
| 22 | 19881 | ICD-9 |
| 23 | 19882 | ICD-9 |
| 24 | 19889 | ICD-9 |
| 25 | 1989 | ICD-9 |
| 26 | 1990 | ICD-9 |
| 27 | 1991 | ICD-9 |
| 28 | 1992 | ICD-9 |
| 29 | C790 | ICD-10 |
| 30 | C7900 | ICD-10 |
| 31 | C7901 | ICD-10 |

| # | ICD Code | Version |
|---|---|---|
| 32 | C7902 | ICD-10 |
| 33 | C791 | ICD-10 |
| 34 | C7910 | ICD-10 |
| 35 | C7911 | ICD-10 |
| 36 | C7919 | ICD-10 |
| 37 | C792 | ICD-10 |
| 38 | C793 | ICD-10 |
| 39 | C7931 | ICD-10 |
| 40 | C7932 | ICD-10 |
| 41 | C794 | ICD-10 |
| 42 | C7940 | ICD-10 |
| 43 | C7949 | ICD-10 |
| 44 | C795 | ICD-10 |
| 45 | C7951 | ICD-10 |
| 46 | C7952 | ICD-10 |
| 47 | C796 | ICD-10 |
| 48 | C7960 | ICD-10 |
| 49 | C7961 | ICD-10 |
| 50 | C7962 | ICD-10 |
| 51 | C7963 | ICD-10 |
| 52 | C797 | ICD-10 |
| 53 | C7970 | ICD-10 |
| 54 | C7971 | ICD-10 |
| 55 | C7972 | ICD-10 |
| 56 | C798 | ICD-10 |
| 57 | C7981 | ICD-10 |
| 58 | C7982 | ICD-10 |
| 59 | C7989 | ICD-10 |
| 60 | C799 | ICD-10 |
| 61 | C780 | ICD-10 |
| 62 | C7800 | ICD-10 |
| 63 | C7801 | ICD-10 |
| 64 | C7802 | ICD-10 |
| 65 | C781 | ICD-10 |
| 66 | C782 | ICD-10 |
| 67 | C783 | ICD-10 |
| 68 | C7830 | ICD-10 |
| 69 | C7839 | ICD-10 |
| 70 | C784 | ICD-10 |
| 71 | C785 | ICD-10 |
| 72 | C786 | ICD-10 |
| 73 | C787 | ICD-10 |
| 74 | C788 | ICD-10 |
| 75 | C7880 | ICD-10 |
| 76 | C7889 | ICD-10 |
| 77 | C770 | ICD-10 |
| 78 | C771 | ICD-10 |
| 79 | C772 | ICD-10 |
| 80 | C773 | ICD-10 |
| 81 | C774 | ICD-10 |
| 82 | C775 | ICD-10 |
| 83 | C776 | ICD-10 |

| # | ICD Code | Version |
|---|---|---|
| 84 | C777 | ICD-10 |
| 85 | C778 | ICD-10 |
| 86 | C779 | ICD-10 |

Obesity

11 codes

| # | ICD Code | Version |
|---|---|---|
| 1 | 27800 | ICD-9 |
| 2 | 27801 | ICD-9 |
| 3 | E6601 | ICD-10 |
| 4 | E6602 | ICD-10 |
| 5 | E6609 | ICD-10 |
| 6 | E661 | ICD-10 |
| 7 | E662 | ICD-10 |
| 8 | E663 | ICD-10 |
| 9 | E664 | ICD-10 |
| 10 | E668 | ICD-10 |
| 11 | E669 | ICD-10 |

Rheumatoid Arthritis

422 codes

| # | ICD Code | Version |
|---|---|---|
| 1 | 714 | ICD-9 |
| 2 | 7140 | ICD-9 |
| 3 | 7141 | ICD-9 |
| 4 | 7142 | ICD-9 |
| 5 | 7148 | ICD-9 |
| 6 | 71481 | ICD-9 |
| 7 | 71489 | ICD-9 |
| 8 | 7149 | ICD-9 |
| 9 | M06 | ICD-10 |
| 10 | M060 | ICD-10 |
| 11 | M0600 | ICD-10 |
| 12 | M0601 | ICD-10 |
| 13 | M06011 | ICD-10 |
| 14 | M06012 | ICD-10 |
| 15 | M06019 | ICD-10 |
| 16 | M0602 | ICD-10 |
| 17 | M06021 | ICD-10 |
| 18 | M06022 | ICD-10 |
| 19 | M06029 | ICD-10 |
| 20 | M0603 | ICD-10 |
| 21 | M06031 | ICD-10 |
| 22 | M06032 | ICD-10 |
| 23 | M06039 | ICD-10 |
| 24 | M0604 | ICD-10 |

| # | ICD Code | Version |
|---|---|---|
| 25 | M06041 | ICD-10 |
| 26 | M06042 | ICD-10 |
| 27 | M06049 | ICD-10 |
| 28 | M0605 | ICD-10 |
| 29 | M06051 | ICD-10 |
| 30 | M06052 | ICD-10 |
| 31 | M06059 | ICD-10 |
| 32 | M0606 | ICD-10 |
| 33 | M06061 | ICD-10 |
| 34 | M06062 | ICD-10 |
| 35 | M06069 | ICD-10 |
| 36 | M0607 | ICD-10 |
| 37 | M06071 | ICD-10 |
| 38 | M06072 | ICD-10 |
| 39 | M06079 | ICD-10 |
| 40 | M0608 | ICD-10 |
| 41 | M0609 | ICD-10 |
| 42 | M060A | ICD-10 |
| 43 | M061 | ICD-10 |
| 44 | M062 | ICD-10 |
| 45 | M0620 | ICD-10 |
| 46 | M0621 | ICD-10 |
| 47 | M06211 | ICD-10 |
| 48 | M06212 | ICD-10 |
| 49 | M06219 | ICD-10 |
| 50 | M0622 | ICD-10 |
| 51 | M06221 | ICD-10 |
| 52 | M06222 | ICD-10 |
| 53 | M06229 | ICD-10 |
| 54 | M0623 | ICD-10 |
| 55 | M06231 | ICD-10 |
| 56 | M06232 | ICD-10 |
| 57 | M06239 | ICD-10 |
| 58 | M0624 | ICD-10 |
| 59 | M06241 | ICD-10 |
| 60 | M06242 | ICD-10 |
| 61 | M06249 | ICD-10 |
| 62 | M0625 | ICD-10 |
| 63 | M06251 | ICD-10 |
| 64 | M06252 | ICD-10 |
| 65 | M06259 | ICD-10 |
| 66 | M0626 | ICD-10 |
| 67 | M06261 | ICD-10 |
| 68 | M06262 | ICD-10 |
| 69 | M06269 | ICD-10 |
| 70 | M0627 | ICD-10 |
| 71 | M06271 | ICD-10 |
| 72 | M06272 | ICD-10 |
| 73 | M06279 | ICD-10 |
| 74 | M0628 | ICD-10 |
| 75 | M0629 | ICD-10 |
| 76 | M063 | ICD-10 |

| # | ICD Code | Version |
|---|---|---|
| 77 | M0630 | ICD-10 |
| 78 | M0631 | ICD-10 |
| 79 | M06311 | ICD-10 |
| 80 | M06312 | ICD-10 |
| 81 | M06319 | ICD-10 |
| 82 | M0632 | ICD-10 |
| 83 | M06321 | ICD-10 |
| 84 | M06322 | ICD-10 |
| 85 | M06329 | ICD-10 |
| 86 | M0633 | ICD-10 |
| 87 | M06331 | ICD-10 |
| 88 | M06332 | ICD-10 |
| 89 | M06339 | ICD-10 |
| 90 | M0634 | ICD-10 |
| 91 | M06341 | ICD-10 |
| 92 | M06342 | ICD-10 |
| 93 | M06349 | ICD-10 |
| 94 | M0635 | ICD-10 |
| 95 | M06351 | ICD-10 |
| 96 | M06352 | ICD-10 |
| 97 | M06359 | ICD-10 |
| 98 | M0636 | ICD-10 |
| 99 | M06361 | ICD-10 |
| 100 | M06362 | ICD-10 |
| 101 | M06369 | ICD-10 |
| 102 | M0637 | ICD-10 |
| 103 | M06371 | ICD-10 |
| 104 | M06372 | ICD-10 |
| 105 | M06379 | ICD-10 |
| 106 | M0638 | ICD-10 |
| 107 | M0639 | ICD-10 |
| 108 | M064 | ICD-10 |
| 109 | M068 | ICD-10 |
| 110 | M0680 | ICD-10 |
| 111 | M0681 | ICD-10 |
| 112 | M06811 | ICD-10 |
| 113 | M06812 | ICD-10 |
| 114 | M06819 | ICD-10 |
| 115 | M0682 | ICD-10 |
| 116 | M06821 | ICD-10 |
| 117 | M06822 | ICD-10 |
| 118 | M06829 | ICD-10 |
| 119 | M0683 | ICD-10 |
| 120 | M06831 | ICD-10 |
| 121 | M06832 | ICD-10 |
| 122 | M06839 | ICD-10 |
| 123 | M0684 | ICD-10 |
| 124 | M06841 | ICD-10 |
| 125 | M06842 | ICD-10 |
| 126 | M06849 | ICD-10 |
| 127 | M0685 | ICD-10 |
| 128 | M06851 | ICD-10 |

| # | ICD Code | Version |
|---|---|---|
| 129 | M06852 | ICD-10 |
| 130 | M06859 | ICD-10 |
| 131 | M0686 | ICD-10 |
| 132 | M06861 | ICD-10 |
| 133 | M06862 | ICD-10 |
| 134 | M06869 | ICD-10 |
| 135 | M0687 | ICD-10 |
| 136 | M06871 | ICD-10 |
| 137 | M06872 | ICD-10 |
| 138 | M06879 | ICD-10 |
| 139 | M0688 | ICD-10 |
| 140 | M0689 | ICD-10 |
| 141 | M05 | ICD-10 |
| 142 | M050 | ICD-10 |
| 143 | M0500 | ICD-10 |
| 144 | M0501 | ICD-10 |
| 145 | M05011 | ICD-10 |
| 146 | M05012 | ICD-10 |
| 147 | M05019 | ICD-10 |
| 148 | M0502 | ICD-10 |
| 149 | M05021 | ICD-10 |
| 150 | M05022 | ICD-10 |
| 151 | M05029 | ICD-10 |
| 152 | M0503 | ICD-10 |
| 153 | M05031 | ICD-10 |
| 154 | M05032 | ICD-10 |
| 155 | M05039 | ICD-10 |
| 156 | M0504 | ICD-10 |
| 157 | M05041 | ICD-10 |
| 158 | M05042 | ICD-10 |
| 159 | M05049 | ICD-10 |
| 160 | M0505 | ICD-10 |
| 161 | M05051 | ICD-10 |
| 162 | M05052 | ICD-10 |
| 163 | M05059 | ICD-10 |
| 164 | M0506 | ICD-10 |
| 165 | M05061 | ICD-10 |
| 166 | M05062 | ICD-10 |
| 167 | M05069 | ICD-10 |
| 168 | M0507 | ICD-10 |
| 169 | M05071 | ICD-10 |
| 170 | M05072 | ICD-10 |
| 171 | M05079 | ICD-10 |
| 172 | M0509 | ICD-10 |
| 173 | M051 | ICD-10 |
| 174 | M0510 | ICD-10 |
| 175 | M0511 | ICD-10 |
| 176 | M05111 | ICD-10 |
| 177 | M05112 | ICD-10 |
| 178 | M05119 | ICD-10 |
| 179 | M0512 | ICD-10 |
| 180 | M05121 | ICD-10 |

| # | ICD Code | Version |
|---|---|---|
| 181 | M05122 | ICD-10 |
| 182 | M05129 | ICD-10 |
| 183 | M0513 | ICD-10 |
| 184 | M05131 | ICD-10 |
| 185 | M05132 | ICD-10 |
| 186 | M05139 | ICD-10 |
| 187 | M0514 | ICD-10 |
| 188 | M05141 | ICD-10 |
| 189 | M05142 | ICD-10 |
| 190 | M05149 | ICD-10 |
| 191 | M0515 | ICD-10 |
| 192 | M05151 | ICD-10 |
| 193 | M05152 | ICD-10 |
| 194 | M05159 | ICD-10 |
| 195 | M0516 | ICD-10 |
| 196 | M05161 | ICD-10 |
| 197 | M05162 | ICD-10 |
| 198 | M05169 | ICD-10 |
| 199 | M0517 | ICD-10 |
| 200 | M05171 | ICD-10 |
| 201 | M05172 | ICD-10 |
| 202 | M05179 | ICD-10 |
| 203 | M0519 | ICD-10 |
| 204 | M052 | ICD-10 |
| 205 | M0520 | ICD-10 |
| 206 | M0521 | ICD-10 |
| 207 | M05211 | ICD-10 |
| 208 | M05212 | ICD-10 |
| 209 | M05219 | ICD-10 |
| 210 | M0522 | ICD-10 |
| 211 | M05221 | ICD-10 |
| 212 | M05222 | ICD-10 |
| 213 | M05229 | ICD-10 |
| 214 | M0523 | ICD-10 |
| 215 | M05231 | ICD-10 |
| 216 | M05232 | ICD-10 |
| 217 | M05239 | ICD-10 |
| 218 | M0524 | ICD-10 |
| 219 | M05241 | ICD-10 |
| 220 | M05242 | ICD-10 |
| 221 | M05249 | ICD-10 |
| 222 | M0525 | ICD-10 |
| 223 | M05251 | ICD-10 |
| 224 | M05252 | ICD-10 |
| 225 | M05259 | ICD-10 |
| 226 | M0526 | ICD-10 |
| 227 | M05261 | ICD-10 |
| 228 | M05262 | ICD-10 |
| 229 | M05269 | ICD-10 |
| 230 | M0527 | ICD-10 |
| 231 | M05271 | ICD-10 |
| 232 | M05272 | ICD-10 |

| # | ICD Code | Version |
|---|---|---|
| 233 | M05279 | ICD-10 |
| 234 | M0529 | ICD-10 |
| 235 | M053 | ICD-10 |
| 236 | M0530 | ICD-10 |
| 237 | M0531 | ICD-10 |
| 238 | M05311 | ICD-10 |
| 239 | M05312 | ICD-10 |
| 240 | M05319 | ICD-10 |
| 241 | M0532 | ICD-10 |
| 242 | M05321 | ICD-10 |
| 243 | M05322 | ICD-10 |
| 244 | M05329 | ICD-10 |
| 245 | M0533 | ICD-10 |
| 246 | M05331 | ICD-10 |
| 247 | M05332 | ICD-10 |
| 248 | M05339 | ICD-10 |
| 249 | M0534 | ICD-10 |
| 250 | M05341 | ICD-10 |
| 251 | M05342 | ICD-10 |
| 252 | M05349 | ICD-10 |
| 253 | M0535 | ICD-10 |
| 254 | M05351 | ICD-10 |
| 255 | M05352 | ICD-10 |
| 256 | M05359 | ICD-10 |
| 257 | M0536 | ICD-10 |
| 258 | M05361 | ICD-10 |
| 259 | M05362 | ICD-10 |
| 260 | M05369 | ICD-10 |
| 261 | M0537 | ICD-10 |
| 262 | M05371 | ICD-10 |
| 263 | M05372 | ICD-10 |
| 264 | M05379 | ICD-10 |
| 265 | M0539 | ICD-10 |
| 266 | M054 | ICD-10 |
| 267 | M0540 | ICD-10 |
| 268 | M0541 | ICD-10 |
| 269 | M05411 | ICD-10 |
| 270 | M05412 | ICD-10 |
| 271 | M05419 | ICD-10 |
| 272 | M0542 | ICD-10 |
| 273 | M05421 | ICD-10 |
| 274 | M05422 | ICD-10 |
| 275 | M05429 | ICD-10 |
| 276 | M0543 | ICD-10 |
| 277 | M05431 | ICD-10 |
| 278 | M05432 | ICD-10 |
| 279 | M05439 | ICD-10 |
| 280 | M0544 | ICD-10 |
| 281 | M05441 | ICD-10 |
| 282 | M05442 | ICD-10 |
| 283 | M05449 | ICD-10 |
| 284 | M0545 | ICD-10 |

| # | ICD Code | Version |
|---|---|---|
| 285 | M05451 | ICD-10 |
| 286 | M05452 | ICD-10 |
| 287 | M05459 | ICD-10 |
| 288 | M0546 | ICD-10 |
| 289 | M05461 | ICD-10 |
| 290 | M05462 | ICD-10 |
| 291 | M05469 | ICD-10 |
| 292 | M0547 | ICD-10 |
| 293 | M05471 | ICD-10 |
| 294 | M05472 | ICD-10 |
| 295 | M05479 | ICD-10 |
| 296 | M0549 | ICD-10 |
| 297 | M055 | ICD-10 |
| 298 | M0550 | ICD-10 |
| 299 | M0551 | ICD-10 |
| 300 | M05511 | ICD-10 |
| 301 | M05512 | ICD-10 |
| 302 | M05519 | ICD-10 |
| 303 | M0552 | ICD-10 |
| 304 | M05521 | ICD-10 |
| 305 | M05522 | ICD-10 |
| 306 | M05529 | ICD-10 |
| 307 | M0553 | ICD-10 |
| 308 | M05531 | ICD-10 |
| 309 | M05532 | ICD-10 |
| 310 | M05539 | ICD-10 |
| 311 | M0554 | ICD-10 |
| 312 | M05541 | ICD-10 |
| 313 | M05542 | ICD-10 |
| 314 | M05549 | ICD-10 |
| 315 | M0555 | ICD-10 |
| 316 | M05551 | ICD-10 |
| 317 | M05552 | ICD-10 |
| 318 | M05559 | ICD-10 |
| 319 | M0556 | ICD-10 |
| 320 | M05561 | ICD-10 |
| 321 | M05562 | ICD-10 |
| 322 | M05569 | ICD-10 |
| 323 | M0557 | ICD-10 |
| 324 | M05571 | ICD-10 |
| 325 | M05572 | ICD-10 |
| 326 | M05579 | ICD-10 |
| 327 | M0559 | ICD-10 |
| 328 | M059 | ICD-10 |
| 329 | M05A | ICD-10 |
| 330 | M056 | ICD-10 |
| 331 | M0560 | ICD-10 |
| 332 | M0561 | ICD-10 |
| 333 | M05611 | ICD-10 |
| 334 | M05612 | ICD-10 |
| 335 | M05619 | ICD-10 |
| 336 | M0562 | ICD-10 |

| # | ICD Code | Version |
|---|---|---|
| 337 | M05621 | ICD-10 |
| 338 | M05622 | ICD-10 |
| 339 | M05629 | ICD-10 |
| 340 | M0563 | ICD-10 |
| 341 | M05631 | ICD-10 |
| 342 | M05632 | ICD-10 |
| 343 | M05639 | ICD-10 |
| 344 | M0564 | ICD-10 |
| 345 | M05641 | ICD-10 |
| 346 | M05642 | ICD-10 |
| 347 | M05649 | ICD-10 |
| 348 | M0565 | ICD-10 |
| 349 | M05651 | ICD-10 |
| 350 | M05652 | ICD-10 |
| 351 | M05659 | ICD-10 |
| 352 | M0566 | ICD-10 |
| 353 | M05661 | ICD-10 |
| 354 | M05662 | ICD-10 |
| 355 | M05669 | ICD-10 |
| 356 | M0567 | ICD-10 |
| 357 | M05671 | ICD-10 |
| 358 | M05672 | ICD-10 |
| 359 | M05679 | ICD-10 |
| 360 | M0569 | ICD-10 |
| 361 | M057 | ICD-10 |
| 362 | M0570 | ICD-10 |
| 363 | M0571 | ICD-10 |
| 364 | M05711 | ICD-10 |
| 365 | M05712 | ICD-10 |
| 366 | M05719 | ICD-10 |
| 367 | M0572 | ICD-10 |
| 368 | M05721 | ICD-10 |
| 369 | M05722 | ICD-10 |
| 370 | M05729 | ICD-10 |
| 371 | M0573 | ICD-10 |
| 372 | M05731 | ICD-10 |
| 373 | M05732 | ICD-10 |
| 374 | M05739 | ICD-10 |
| 375 | M0574 | ICD-10 |
| 376 | M05741 | ICD-10 |
| 377 | M05742 | ICD-10 |
| 378 | M05749 | ICD-10 |
| 379 | M0575 | ICD-10 |
| 380 | M05751 | ICD-10 |
| 381 | M05752 | ICD-10 |
| 382 | M05759 | ICD-10 |
| 383 | M0576 | ICD-10 |
| 384 | M05761 | ICD-10 |
| 385 | M05762 | ICD-10 |
| 386 | M05769 | ICD-10 |
| 387 | M0577 | ICD-10 |
| 388 | M05771 | ICD-10 |

| # | ICD Code | Version |
|---|---|---|
| 389 | M05772 | ICD-10 |
| 390 | M05779 | ICD-10 |
| 391 | M0579 | ICD-10 |
| 392 | M058 | ICD-10 |
| 393 | M0580 | ICD-10 |
| 394 | M0581 | ICD-10 |
| 395 | M05811 | ICD-10 |
| 396 | M05812 | ICD-10 |
| 397 | M05819 | ICD-10 |
| 398 | M0582 | ICD-10 |
| 399 | M05821 | ICD-10 |
| 400 | M05822 | ICD-10 |
| 401 | M05829 | ICD-10 |
| 402 | M0583 | ICD-10 |
| 403 | M05831 | ICD-10 |
| 404 | M05832 | ICD-10 |
| 405 | M05839 | ICD-10 |
| 406 | M0584 | ICD-10 |
| 407 | M05841 | ICD-10 |
| 408 | M05842 | ICD-10 |
| 409 | M05849 | ICD-10 |
| 410 | M0585 | ICD-10 |
| 411 | M05851 | ICD-10 |
| 412 | M05852 | ICD-10 |
| 413 | M05859 | ICD-10 |
| 414 | M0586 | ICD-10 |
| 415 | M05861 | ICD-10 |
| 416 | M05862 | ICD-10 |
| 417 | M05869 | ICD-10 |
| 418 | M0587 | ICD-10 |
| 419 | M05871 | ICD-10 |
| 420 | M05872 | ICD-10 |
| 421 | M05879 | ICD-10 |
| 422 | M0589 | ICD-10 |

Alcohol Abuse / Use Disorder

78 codes

| # | ICD Code | Version |
|---|---|---|
| 1 | 3050 | ICD-9 |
| 2 | 30500 | ICD-9 |
| 3 | 30501 | ICD-9 |
| 4 | 30502 | ICD-9 |
| 5 | 30503 | ICD-9 |
| 6 | F101 | ICD-10 |
| 7 | F1010 | ICD-10 |
| 8 | F1011 | ICD-10 |
| 9 | F1012 | ICD-10 |
| 10 | F10120 | ICD-10 |
| 11 | F10121 | ICD-10 |

| # | ICD Code | Version |
|---|---|---|
| 12 | F10129 | ICD-10 |
| 13 | F1013 | ICD-10 |
| 14 | F10130 | ICD-10 |
| 15 | F10131 | ICD-10 |
| 16 | F10132 | ICD-10 |
| 17 | F10139 | ICD-10 |
| 18 | F1014 | ICD-10 |
| 19 | F1015 | ICD-10 |
| 20 | F10150 | ICD-10 |
| 21 | F10151 | ICD-10 |
| 22 | F10159 | ICD-10 |
| 23 | F1018 | ICD-10 |
| 24 | F10180 | ICD-10 |
| 25 | F10181 | ICD-10 |
| 26 | F10182 | ICD-10 |
| 27 | F10188 | ICD-10 |
| 28 | F1019 | ICD-10 |
| 29 | F102 | ICD-10 |
| 30 | F1020 | ICD-10 |
| 31 | F1021 | ICD-10 |
| 32 | F1022 | ICD-10 |
| 33 | F10220 | ICD-10 |
| 34 | F10221 | ICD-10 |
| 35 | F10229 | ICD-10 |
| 36 | F1023 | ICD-10 |
| 37 | F10230 | ICD-10 |
| 38 | F10231 | ICD-10 |
| 39 | F10232 | ICD-10 |
| 40 | F10239 | ICD-10 |
| 41 | F1024 | ICD-10 |
| 42 | F1025 | ICD-10 |
| 43 | F10250 | ICD-10 |
| 44 | F10251 | ICD-10 |
| 45 | F10259 | ICD-10 |
| 46 | F1026 | ICD-10 |
| 47 | F1027 | ICD-10 |
| 48 | F1028 | ICD-10 |
| 49 | F10280 | ICD-10 |
| 50 | F10281 | ICD-10 |
| 51 | F10282 | ICD-10 |
| 52 | F10288 | ICD-10 |
| 53 | F1029 | ICD-10 |
| 54 | F109 | ICD-10 |
| 55 | F1090 | ICD-10 |
| 56 | F1091 | ICD-10 |
| 57 | F1092 | ICD-10 |
| 58 | F10920 | ICD-10 |
| 59 | F10921 | ICD-10 |
| 60 | F10929 | ICD-10 |
| 61 | F1093 | ICD-10 |
| 62 | F10930 | ICD-10 |
| 63 | F10931 | ICD-10 |

| # | ICD Code | Version |
|---|---|---|
| 64 | F10932 | ICD-10 |
| 65 | F10939 | ICD-10 |
| 66 | F1094 | ICD-10 |
| 67 | F1095 | ICD-10 |
| 68 | F10950 | ICD-10 |
| 69 | F10951 | ICD-10 |
| 70 | F10959 | ICD-10 |
| 71 | F1096 | ICD-10 |
| 72 | F1097 | ICD-10 |
| 73 | F1098 | ICD-10 |
| 74 | F10980 | ICD-10 |
| 75 | F10981 | ICD-10 |
| 76 | F10982 | ICD-10 |
| 77 | F10988 | ICD-10 |
| 78 | F1099 | ICD-10 |

Venous Thromboembolism (VTE) – Complication

174 codes

| # | ICD Code | Version |
|---|---|---|
| 1 | 415 | ICD-9 |
| 2 | 4150 | ICD-9 |
| 3 | 4151 | ICD-9 |
| 4 | 41511 | ICD-9 |
| 5 | 41512 | ICD-9 |
| 6 | 41513 | ICD-9 |
| 7 | 41519 | ICD-9 |
| 8 | 416 | ICD-9 |
| 9 | 4160 | ICD-9 |
| 10 | 4161 | ICD-9 |
| 11 | 4162 | ICD-9 |
| 12 | 4168 | ICD-9 |
| 13 | 4169 | ICD-9 |
| 14 | 451 | ICD-9 |
| 15 | 4510 | ICD-9 |
| 16 | 4511 | ICD-9 |
| 17 | 45119 | ICD-9 |
| 18 | 4512 | ICD-9 |
| 19 | 4518 | ICD-9 |
| 20 | 45181 | ICD-9 |
| 21 | 45182 | ICD-9 |
| 22 | 45183 | ICD-9 |
| 23 | 45184 | ICD-9 |
| 24 | 45189 | ICD-9 |
| 25 | 4519 | ICD-9 |
| 26 | 453 | ICD-9 |
| 27 | 4530 | ICD-9 |
| 28 | 4531 | ICD-9 |
| 29 | 4532 | ICD-9 |
| 30 | 4533 | ICD-9 |

| # | ICD Code | Version |
|---|---|---|
| 31 | 4534 | ICD-9 |
| 32 | 45340 | ICD-9 |
| 33 | 45341 | ICD-9 |
| 34 | 45342 | ICD-9 |
| 35 | 4535 | ICD-9 |
| 36 | 45350 | ICD-9 |
| 37 | 45351 | ICD-9 |
| 38 | 45352 | ICD-9 |
| 39 | 4536 | ICD-9 |
| 40 | 4537 | ICD-9 |
| 41 | 45371 | ICD-9 |
| 42 | 45372 | ICD-9 |
| 43 | 45373 | ICD-9 |
| 44 | 45374 | ICD-9 |
| 45 | 45375 | ICD-9 |
| 46 | 45376 | ICD-9 |
| 47 | 45377 | ICD-9 |
| 48 | 45379 | ICD-9 |
| 49 | 4538 | ICD-9 |
| 50 | 45381 | ICD-9 |
| 51 | 45382 | ICD-9 |
| 52 | 45383 | ICD-9 |
| 53 | 45384 | ICD-9 |
| 54 | 45385 | ICD-9 |
| 55 | 45386 | ICD-9 |
| 56 | 45387 | ICD-9 |
| 57 | 45389 | ICD-9 |
| 58 | 4539 | ICD-9 |
| 59 | I80 | ICD-10 |
| 60 | I800 | ICD-10 |
| 61 | I8000 | ICD-10 |
| 62 | I8001 | ICD-10 |
| 63 | I8002 | ICD-10 |
| 64 | I8003 | ICD-10 |
| 65 | I801 | ICD-10 |
| 66 | I8010 | ICD-10 |
| 67 | I8011 | ICD-10 |
| 68 | I8012 | ICD-10 |
| 69 | I8013 | ICD-10 |
| 70 | I802 | ICD-10 |
| 71 | I8020 | ICD-10 |
| 72 | I80201 | ICD-10 |
| 73 | I80202 | ICD-10 |
| 74 | I80203 | ICD-10 |
| 75 | I80209 | ICD-10 |
| 76 | I8021 | ICD-10 |
| 77 | I80211 | ICD-10 |
| 78 | I80212 | ICD-10 |
| 79 | I80213 | ICD-10 |
| 80 | I80219 | ICD-10 |
| 81 | I8022 | ICD-10 |
| 82 | I80221 | ICD-10 |

| # | ICD Code | Version |
|---|---|---|
| 83 | I80222 | ICD-10 |
| 84 | I80223 | ICD-10 |
| 85 | I80229 | ICD-10 |
| 86 | I8023 | ICD-10 |
| 87 | I80231 | ICD-10 |
| 88 | I80232 | ICD-10 |
| 89 | I80233 | ICD-10 |
| 90 | I80239 | ICD-10 |
| 91 | I8024 | ICD-10 |
| 92 | I80241 | ICD-10 |
| 93 | I80242 | ICD-10 |
| 94 | I80243 | ICD-10 |
| 95 | I80249 | ICD-10 |
| 96 | I8025 | ICD-10 |
| 97 | I80251 | ICD-10 |
| 98 | I80252 | ICD-10 |
| 99 | I80253 | ICD-10 |
| 100 | I80259 | ICD-10 |
| 101 | I8029 | ICD-10 |
| 102 | I80291 | ICD-10 |
| 103 | I80292 | ICD-10 |
| 104 | I80293 | ICD-10 |
| 105 | I80299 | ICD-10 |
| 106 | I803 | ICD-10 |
| 107 | I808 | ICD-10 |
| 108 | I809 | ICD-10 |
| 109 | I82 | ICD-10 |
| 110 | I820 | ICD-10 |
| 111 | I821 | ICD-10 |
| 112 | I822 | ICD-10 |
| 113 | I8221 | ICD-10 |
| 114 | I8222 | ICD-10 |
| 115 | I8223 | ICD-10 |
| 116 | I8229 | ICD-10 |
| 117 | I823 | ICD-10 |
| 118 | I8231 | ICD-10 |
| 119 | I8232 | ICD-10 |
| 120 | I8233 | ICD-10 |
| 121 | I8239 | ICD-10 |
| 122 | I824 | ICD-10 |
| 123 | I8241 | ICD-10 |
| 124 | I8242 | ICD-10 |
| 125 | I8243 | ICD-10 |
| 126 | I8249 | ICD-10 |
| 127 | I825 | ICD-10 |
| 128 | I8251 | ICD-10 |
| 129 | I8252 | ICD-10 |
| 130 | I8253 | ICD-10 |
| 131 | I8259 | ICD-10 |
| 132 | I826 | ICD-10 |
| 133 | I8261 | ICD-10 |
| 134 | I8262 | ICD-10 |

| # | ICD Code | Version |
|---|---|---|
| 135 | I8263 | ICD-10 |
| 136 | I8269 | ICD-10 |
| 137 | I829 | ICD-10 |
| 138 | I8291 | ICD-10 |
| 139 | I8292 | ICD-10 |
| 140 | I8293 | ICD-10 |
| 141 | I8299 | ICD-10 |
| 142 | I82A | ICD-10 |
| 143 | I82A1 | ICD-10 |
| 144 | I82A2 | ICD-10 |
| 145 | I82A3 | ICD-10 |
| 146 | I82A9 | ICD-10 |
| 147 | I82B | ICD-10 |
| 148 | I82B1 | ICD-10 |
| 149 | I82B2 | ICD-10 |
| 150 | I82B3 | ICD-10 |
| 151 | I82B9 | ICD-10 |
| 152 | I82C | ICD-10 |
| 153 | I82C1 | ICD-10 |
| 154 | I82C2 | ICD-10 |
| 155 | I82C3 | ICD-10 |
| 156 | I82C9 | ICD-10 |
| 157 | I828 | ICD-10 |
| 158 | I8281 | ICD-10 |
| 159 | I8282 | ICD-10 |
| 160 | I8283 | ICD-10 |
| 161 | I8289 | ICD-10 |
| 162 | I8290 | ICD-10 |
| 163 | I26 | ICD-10 |
| 164 | I260 | ICD-10 |
| 165 | I2600 | ICD-10 |
| 166 | I26001 | ICD-10 |
| 167 | I26002 | ICD-10 |
| 168 | I26009 | ICD-10 |
| 169 | I2609 | ICD-10 |
| 170 | I26090 | ICD-10 |
| 171 | I26092 | ICD-10 |
| 172 | I26093 | ICD-10 |
| 173 | I26094 | ICD-10 |
| 174 | I26099 | ICD-10 |

Venous Thromboembolism (VTE) – Past History

174 codes

| # | ICD Code | Version |
|---|---|---|
| 1 | 415 | ICD-9 |
| 2 | 4150 | ICD-9 |
| 3 | 4151 | ICD-9 |
| 4 | 41511 | ICD-9 |
| 5 | 41512 | ICD-9 |

| # | ICD Code | Version |
|---|---|---|
| 6 | 41513 | ICD-9 |
| 7 | 41519 | ICD-9 |
| 8 | 416 | ICD-9 |
| 9 | 4160 | ICD-9 |
| 10 | 4161 | ICD-9 |
| 11 | 4162 | ICD-9 |
| 12 | 4168 | ICD-9 |
| 13 | 4169 | ICD-9 |
| 14 | 451 | ICD-9 |
| 15 | 4510 | ICD-9 |
| 16 | 4511 | ICD-9 |
| 17 | 45119 | ICD-9 |
| 18 | 4512 | ICD-9 |
| 19 | 4518 | ICD-9 |
| 20 | 45181 | ICD-9 |
| 21 | 45182 | ICD-9 |
| 22 | 45183 | ICD-9 |
| 23 | 45184 | ICD-9 |
| 24 | 45189 | ICD-9 |
| 25 | 4519 | ICD-9 |
| 26 | 453 | ICD-9 |
| 27 | 4530 | ICD-9 |
| 28 | 4531 | ICD-9 |
| 29 | 4532 | ICD-9 |
| 30 | 4533 | ICD-9 |
| 31 | 4534 | ICD-9 |
| 32 | 45340 | ICD-9 |
| 33 | 45341 | ICD-9 |
| 34 | 45342 | ICD-9 |
| 35 | 4535 | ICD-9 |
| 36 | 45350 | ICD-9 |
| 37 | 45351 | ICD-9 |
| 38 | 45352 | ICD-9 |
| 39 | 4536 | ICD-9 |
| 40 | 4537 | ICD-9 |
| 41 | 45371 | ICD-9 |
| 42 | 45372 | ICD-9 |
| 43 | 45373 | ICD-9 |
| 44 | 45374 | ICD-9 |
| 45 | 45375 | ICD-9 |
| 46 | 45376 | ICD-9 |
| 47 | 45377 | ICD-9 |
| 48 | 45379 | ICD-9 |
| 49 | 4538 | ICD-9 |
| 50 | 45381 | ICD-9 |
| 51 | 45382 | ICD-9 |
| 52 | 45383 | ICD-9 |
| 53 | 45384 | ICD-9 |
| 54 | 45385 | ICD-9 |
| 55 | 45386 | ICD-9 |
| 56 | 45387 | ICD-9 |
| 57 | 45389 | ICD-9 |

| # | ICD Code | Version |
|---|---|---|
| 58 | 4539 | ICD-9 |
| 59 | I80 | ICD-10 |
| 60 | I800 | ICD-10 |
| 61 | I8000 | ICD-10 |
| 62 | I8001 | ICD-10 |
| 63 | I8002 | ICD-10 |
| 64 | I8003 | ICD-10 |
| 65 | I801 | ICD-10 |
| 66 | I8010 | ICD-10 |
| 67 | I8011 | ICD-10 |
| 68 | I8012 | ICD-10 |
| 69 | I8013 | ICD-10 |
| 70 | I802 | ICD-10 |
| 71 | I8020 | ICD-10 |
| 72 | I80201 | ICD-10 |
| 73 | I80202 | ICD-10 |
| 74 | I80203 | ICD-10 |
| 75 | I80209 | ICD-10 |
| 76 | I8021 | ICD-10 |
| 77 | I80211 | ICD-10 |
| 78 | I80212 | ICD-10 |
| 79 | I80213 | ICD-10 |
| 80 | I80219 | ICD-10 |
| 81 | I8022 | ICD-10 |
| 82 | I80221 | ICD-10 |
| 83 | I80222 | ICD-10 |
| 84 | I80223 | ICD-10 |
| 85 | I80229 | ICD-10 |
| 86 | I8023 | ICD-10 |
| 87 | I80231 | ICD-10 |
| 88 | I80232 | ICD-10 |
| 89 | I80233 | ICD-10 |
| 90 | I80239 | ICD-10 |
| 91 | I8024 | ICD-10 |
| 92 | I80241 | ICD-10 |
| 93 | I80242 | ICD-10 |
| 94 | I80243 | ICD-10 |
| 95 | I80249 | ICD-10 |
| 96 | I8025 | ICD-10 |
| 97 | I80251 | ICD-10 |
| 98 | I80252 | ICD-10 |
| 99 | I80253 | ICD-10 |
| 100 | I80259 | ICD-10 |
| 101 | I8029 | ICD-10 |
| 102 | I80291 | ICD-10 |
| 103 | I80292 | ICD-10 |
| 104 | I80293 | ICD-10 |
| 105 | I80299 | ICD-10 |
| 106 | I803 | ICD-10 |
| 107 | I808 | ICD-10 |
| 108 | I809 | ICD-10 |
| 109 | I82 | ICD-10 |

| # | ICD Code | Version |
|---|---|---|
| 110 | I820 | ICD-10 |
| 111 | I821 | ICD-10 |
| 112 | I822 | ICD-10 |
| 113 | I8221 | ICD-10 |
| 114 | I8222 | ICD-10 |
| 115 | I8223 | ICD-10 |
| 116 | I8229 | ICD-10 |
| 117 | I823 | ICD-10 |
| 118 | I8231 | ICD-10 |
| 119 | I8232 | ICD-10 |
| 120 | I8233 | ICD-10 |
| 121 | I8239 | ICD-10 |
| 122 | I824 | ICD-10 |
| 123 | I8241 | ICD-10 |
| 124 | I8242 | ICD-10 |
| 125 | I8243 | ICD-10 |
| 126 | I8249 | ICD-10 |
| 127 | I825 | ICD-10 |
| 128 | I8251 | ICD-10 |
| 129 | I8252 | ICD-10 |
| 130 | I8253 | ICD-10 |
| 131 | I8259 | ICD-10 |
| 132 | I826 | ICD-10 |
| 133 | I8261 | ICD-10 |
| 134 | I8262 | ICD-10 |
| 135 | I8263 | ICD-10 |
| 136 | I8269 | ICD-10 |
| 137 | I829 | ICD-10 |
| 138 | I8291 | ICD-10 |
| 139 | I8292 | ICD-10 |
| 140 | I8293 | ICD-10 |
| 141 | I8299 | ICD-10 |
| 142 | I82A | ICD-10 |
| 143 | I82A1 | ICD-10 |
| 144 | I82A2 | ICD-10 |
| 145 | I82A3 | ICD-10 |
| 146 | I82A9 | ICD-10 |
| 147 | I82B | ICD-10 |
| 148 | I82B1 | ICD-10 |
| 149 | I82B2 | ICD-10 |
| 150 | I82B3 | ICD-10 |
| 151 | I82B9 | ICD-10 |
| 152 | I82C | ICD-10 |
| 153 | I82C1 | ICD-10 |
| 154 | I82C2 | ICD-10 |
| 155 | I82C3 | ICD-10 |
| 156 | I82C9 | ICD-10 |
| 157 | I828 | ICD-10 |
| 158 | I8281 | ICD-10 |
| 159 | I8282 | ICD-10 |
| 160 | I8283 | ICD-10 |
| 161 | I8289 | ICD-10 |

| # | ICD Code | Version |
|---|---|---|
| 162 | I8290 | ICD-10 |
| 163 | I26 | ICD-10 |
| 164 | I260 | ICD-10 |
| 165 | I2600 | ICD-10 |
| 166 | I26001 | ICD-10 |
| 167 | I26002 | ICD-10 |
| 168 | I26009 | ICD-10 |
| 169 | I2609 | ICD-10 |
| 170 | I26090 | ICD-10 |
| 171 | I26092 | ICD-10 |
| 172 | I26093 | ICD-10 |
| 173 | I26094 | ICD-10 |
| 174 | I26099 | ICD-10 |
