## Supplemental File 2 - TF-IDF Keywords for "MIMIC-IV Phenotype Atlas (MIPA): A Publicly Available Dataset for EHR Phenotyping"

### 1 Supplemental File 2 - TF-IDF Keywords

Supplementary Methods 2. TF-IDF Keyword Lists for Phenotyping

This supplementary section provides the phenotype-specific keyword dictionaries used for the TF-IDF baseline models. Keywords include canonical names, abbreviations, misspellings, and clinical variants commonly found in discharge summaries.

| Phenotype | Keywords |
|---|---|
| C. difficile (complication) | c. diff; cdiff; clostridium difficile; clostridium dificile; clostridium deficile; c diff; c-diff; c difficle; cdif; cdifficile; cdifficil; cdiff infection; cdiff recurrence; cdiff severe; cdiff sepsis; cdiff shock; cdiff toxic; cdiff colectomy; cdiff surgery; cdiff icu; cdiff death; toxic megacolon; pseudomembranous colitis |
| C. difficile (past history) | history of c. diff; hx of c diff; past cdiff; previous clostridium difficile; resolved cdiff; prior cdiff; remote cdiff; cdiff hx; cdiff in past; cdiff previously treated; cdiff (resolved); cdiff (history); cdiff (hx) |
| Dementia | dementia; alzheimer; alzheimers; alzeimer; alzheimr; cognitive decline; memory loss; memory impairment; cognitive impairment; vascular dementia; lewy body; frontotemporal; progressive forgetfulness; disoriented; disorientation; confused; confusion; impaired cognition; impaired memory; delirium superimposed on dementia; senile dementia; neurocognitive disorder; demensia; demntia |
| Depression | depression; depressive disorder; major depression; mood disorder; depressed mood; dysthymia; sadness; anhedonia; low mood; depressive symptoms; mdd; recurrent depression; bipolar depression; postpartum depression; psychotic depression; melancholic depression; suicidal ideation; hopelessness; tearful; loss of interest; depresion; depresion disorder; depresive |
| Type 1 Diabetes | type 1 diabetes; t1dm; juvenile diabetes; insulin dependent diabetes; type i diabetes; autoimmune diabetes; diabetes since childhood; onset in childhood; c-peptide negative; diabetes diagnosed young; insulin pump; diabetes mellitus type 1; brittle diabetes; t1d; type one diabetes; type-1 diabetes; type1 diabetes |
| Type 2 Diabetes | type 2 diabetes; t2dm; non-insulin dependent diabetes; adult onset diabetes; type ii diabetes; diabetes mellitus type 2; insulin resistance; metabolic syndrome; hyperglycemia; oral hypoglycemics; diabetes diagnosed as adult; diabetes with obesity; diabetes with hypertension; t2d; type two diabetes; type-2 diabetes; type2 diabetes |

| Phenotype | Keywords |
| --- | --- |
| Heart Failure with Reduced EF (HFrEF) | hfref; systolic heart failure; reduced ejection fraction; heart failure with reduced ejection fraction; low ef; ef < 40; systolic dysfunction; ischemic cardiomyopathy; dilated cardiomyopathy; chf with reduced ef; lvef reduced; heart failure systolic; hfrEF; hfr ef; hfr-ejection fraction; hfrf; systolic hf; systolic chf |
| Heart Failure with Preserved EF (HFpEF) | hfpef; diastolic heart failure; preserved ejection fraction; heart failure with preserved ejection fraction; ef > 50; diastolic dysfunction; chf with preserved ef; lvef preserved; heart failure diastolic; hfpEF; hfp ef; hfp-ejection fraction; hfp f; diastolic hf; diastolic chf |
| Hypertension | hypertension; high blood pressure; htn; elevated bp; essential hypertension; chronic hypertension; uncontrolled hypertension; hypertensive; bp > 140/90; bp elevated; hypertensive urgency; hypertensive emergency; hbp; htn.; htn: |
| Systemic Lupus (SLE) | sle; lupus; systemic lupus erythematosus; autoimmune lupus; lupus nephritis; lupus flare; lupus anticoagulant; lupus rash; lupus arthritis; lupus cerebritis; lupus serositis; lupus diagnosis; systemic lupus; lupis; lupos |
| Metastatic Cancer | metastatic cancer; metastases; stage iv cancer; secondary cancer; cancer spread; distant metastasis; bone metastasis; liver metastasis; lung metastasis; brain metastasis; mets; metastatic disease; cancer with mets; malignant neoplasm with metastasis; metastatic ca; metastatic carcinoma; metastatic tumour; metastatic tumor; metastatic ca. |
| Obesity | obesity; obese; morbid obesity; bmi > 30; bmi > 35; bmi > 40; severe obesity; weight > 100kg; weight > 120kg; body mass index high; adiposity; overweight; super obesity; obesity hypoventilation; obesety; obes; obesitas |
| Rheumatoid Arthritis | rheumatoid arthritis; ra; inflammatory arthritis; seropositive arthritis; seronegative arthritis; erosive arthritis; rheumatoid factor positive; anti-ccp positive; morning stiffness; joint swelling; joint deformity; synovitis; rheumatoid nodules; rheumatoid vasculitis; rheum arth; rheum arthritis; rheumarth; rheum-arthritis |
| Alcohol Abuse | alcohol abuse; alcohol dependence; alcoholism; etoh abuse; etoh dependence; alcohol use disorder; heavy drinking; binge drinking; alcohol withdrawal; alcohol intoxication; alcoholic liver disease; alcoholic hepatitis; alcoholic cirrhosis; alcoholic pancreatitis; alcoholic neuropathy; etoh; etoh use; etoh intoxication; etoh withdrawal; etoh abuse disorder |

| Phenotype | Keywords |
|---|---|
| Venous Thromboembolism (Complication) | vte; dvt; pe; venous thromboembolism complication; deep vein thrombosis; pulmonary embolism; thromboembolism; venous clot; venous thrombosis; venous embolism; recurrent dvt; recurrent pe; dvt complication; pe complication; vte event; vte recurrence; venous thrombus; venous embolus; venous thromboembolic; venous thromboembolic event |
| Venous Thromboembolism (Past History) | history of vte; past dvt; previous pe; history of venous thromboembolism; prior dvt; prior pe; remote vte; remote dvt; remote pe; resolved dvt; resolved pe; dvt hx; pe hx; vte hx; hx vte; hx dvt; hx pe |
