## Supplemental File 3- Supervised Learning Methods and Large Language Model Pipeline for "MIMIC-IV Phenotype Atlas (MIPA): A Publicly Available Dataset for EHR Phenotyping"

### 1 Supplemental File 3 - Supervised Learning Methods and Large Language Model Pipeline

#### Technical Appendix

#### Supervised Machine Learning

**Data Preprocessing and Quality Control** To minimize the impact of missingness, we applied a two-stage sparsity filtering process for the validation and test sets. Observations with >30% missing values were excluded from validation and test sets. Features missing in >20% of observations were removed.

We handled missing data differently according to the supervised learning model. For logistic regression, we used a KNN imputer with n=7; for both tree-based models ( random forest and gradient boosting), we used median imputation. Imputation was conducted separately on the training and validation sets to avoid information leakage. We did not implement handling of missing values for Naive Bayes, as the model tolerates missing values natively

**Feature Selection and Model Training** Models were trained using the Anchor-and-Learn framework, a weakly semi-supervised algorithm allowing learning using silver labels.

- **Stage 1:** Train an initial model on silver-labeled data using the entire set of candidate features Xfull (~2,000 features) using the training dataset labeled with Y*. From this stage, extract the subset of predictive features Xsel from Xfull using absolute coefficient values (linear models), feature importance (tree-based), or ANOVA F-score (Naive Bayes).
- **Stage 2:** Retrain the same model using Xsel on the validation set using Y. For logistic regression, Elastic-Net regularization was applied to mitigate collinearity. Additionally, all feature counts were normalized by length of stay, and hyperparameters were optimized for F1-score using stratified cross-validation. Tuned hyperparameters included below.
- **Random forest:** number of trees, maximum depth, minimum samples per split.
- **Gradient boosting:** number of boosting stages, depth, learning rate.
- **Naive Bayes:** variance smoothing.

#### LLM Prompting Details

GPT-4o (openai/chatgpt-4o-latest) was queried with discharge summaries using zero-shot prompts (temperature 0.5). No prompt engineering tuning was conducted. The model was prompted to return the classification given the entire discharge summary. The prompts used are provided below. Provider was Open Router, and analysis were conducted in September 2025.

**System Prompt** You are a medical researcher specializing in analyzing discharge summaries for phenotype classification in epidemiologic studies.

Your task is to determine the presence or absence of a given condition based on the available evidence in the discharge summary.

**Guidelines**

- Thoroughly examine the discharge summary for all relevant medical information.
- Prioritize documented clinical evidence over speculation.
- Interpret standard medical abbreviations and terminologies accurately.

**Classification Approach**

- Make decisive classifications based on the strongest available evidence.
- For uncertain cases, favor classifications supported by evidence.
- Maintain high accuracy, as your classifications will directly impact the study's validity.

**User Prompt** Analyze the discharge summary provided below to determine the presence of {phenotype}.

Use a detailed reasoning process as a medical doctor would to arrive at your conclusion.

Provide a JSON object with your final judgment, including a classification status and a justification for your decision.

It is imperative that your response be accurate, as errors cannot be tolerated.

**Discharge Summary** {discharge_summary}

**Step-by-Step Reasoning**

1. Identify and list the key clinical features mentioned in the summary that are relevant to {phenotype}.
2. Assess whether these features align more with the presence or absence of {phenotype}. Only output 1 if the evidence is conclusive.
3. Summarize your findings based on the evidence and make a professional judgment.

**Output Format** Output in JSON format with the following three keys: phenotype, status, and justification.

**DO NOT PROVIDE ANY OTHER SURROUNDING TEXT THAN THE JUSTIFICATION.**

**Output** {{"phenotype": "{phenotype}", "status": "1 or 0", "justification": "Your 50-token Justification Here"}}
