## Supplemental File 4 - ACCORD checklist table for "MIMIC-IV Phenotype Atlas (MIPA): A Publicly Available Dataset for EHR Phenotyping"

### 1 Supplemental File 4 - ACCORD Checklist Table

##### Supplementary Table Sx. Accord Checklist Mapping (MIPA Dataset)

| ACCORD item | Checklist completed? | How this was handled in this paper |
|---|---|---|
| T1 – Title identifies consensus exercise | No | The title does not explicitly identify the study as a consensus exercise; consensus is described in the Methods. |
| I1 – Rationale for choosing consensus | Yes | Consensus adjudication was chosen to resolve annotation disagreements and generate high-confidence gold labels. |
| I2 – Aim, audience, and scope | Yes | Aim was to generate expert-annotated phenotypes for the research community; scope is single-institution. |
| I3 – Update of existing document | Not applicable | |
| M1 – Registration | No | The annotation consensus exercise was not prospectively registered. |
| M2 – Leadership / steering committee | Partially | The study was led by the authors; no formal steering committee structure was used. |
| M3 – Panellist inclusion criteria | Yes | Annotators were clinicians selected for relevant medical expertise; two annotators used for feasibility. |
| M4 – Recruitment process | Not applicable | Annotators were study investigators and not externally recruited. |
| M5 – Public or patient involvement | Not applicable | No patient or public involvement occurred. |
| M6 – Preparatory research | Yes | Phenotype definitions and ICD code lists were curated before annotation. |
| M7 – Systematic literature search | Not applicable | No systematic literature review was conducted for consensus purposes. |
| M8 – Evidence summarisation | Partially | Phenotype definitions were provided, but no formal evidence summaries were distributed. |
| M9 – Consensus method | Yes | Independent annotation followed by joint adjudication discussions. |
| M10 – Presentation of items | Yes | Each discharge summary was reviewed for presence/absence of predefined phenotypes. |

| ACCORD item | Checklist completed? | How this was handled in this paper |
|---|---|---|
| M11 – Objective of each step | Yes | Independent review assessed agreement; adjudication resolved disagreements. |
| M12 – Definition of consensus | Yes | Consensus required agreement after adjudication; highly discordant cases were excluded. |
| M13 – Subsequent voting rounds | Not applicable | No iterative voting rounds were used. |
| M14 – Response collection mode | Yes | Annotations were collected independently via Labelbox, then reviewed jointly. |
| M15 – Processing of responses | Yes | Disagreements were quantified, categorised, and resolved or excluded. |
| M16 – Piloting | No | Annotation materials were not formally piloted. |
| M17 – Feedback to panellists | Not applicable | No iterative feedback rounds were conducted. |
| M18 – Anonymity | No | Annotators were not anonymised during consensus discussions. |
| M19 – Steering committee voting | Not applicable | No steering committee voting occurred. |
| M20 – Incentives | Not applicable | No incentives were provided. |
| M21 – Accessibility adaptations | Not applicable | All work was conducted in English by clinicians fluent in English. |
| R1 – Dates and duration | Partially | Overall annotation counts are reported, but detailed timelines are not. |
| R2 – Protocol deviations | Not applicable | No protocol deviations were reported. |
| R3 – Participant characteristics | Partially | Annotator roles are described |
| R4 – Final consensus outcomes | Yes | Final dataset size and agreement metrics are reported. |
| R5 – Modified or removed items | Yes | Documents were excluded when consensus could not be reached. |
| D1 – Strengths and limitations | Yes | Limitations of the consensus process are discussed. |
| D2 – Consistency with literature | Not applicable | No recommendations are compared with prior consensus literature. |
| O1 – Endorsing organisations | Not applicable | No endorsing organisations were involved. |
| O2 – Conflicts of interest | Yes | Conflicts of interest are disclosed. |
| O3 – Funding and funder role | Yes | Funding sources and funder role are disclosed. |
