## Supplementary Tables (S1, S2) for "MIMIC-IV Phenotype Atlas (MIPA): A Publicly Available Dataset for EHR Phenotyping"

### 1 Supplementary Tables

**Table S1:** Keywords used for TF-IDF for each phenotype

| Phenotype | Keywords (abbreviated list, full list includes typos and other variations) |
|---|---|
| Alcohol Abuse | alcohol abuse, alcohol dependence, alcoholism, etoh abuse, etoh dependence, alcohol use disorder, heavy drinking |
| C.Diff Complication | c. diff, cdiff, clostridium difficile, clostridium dificile, clostridium deficile, c diff, c-diff |
| Past C.Diff | history of c. diff, hx of c diff, past cdiff, previous clostridium difficile, resolved cdiff, prior cdiff, remote cdiff |
| Dementia | dementia, alzheimer, alzheimers, alzeimer, alzheimr, cognitive decline, parkinson, memory loss |
| Depression | depression, depressive disorder, major depression, mood disorder, depressed mood, dysthymia, sadness |
| Type 1 Diabetes | type 1 diabetes, t1dm, juvenile diabetes, insulin dependent diabetes, type i diabetes, autoimmune diabetes, diabetes since childhood |
| Type 2 Diabetes | type 2 diabetes, t2dm, non-insulin dependent diabetes, adult onset diabetes, type ii diabetes, diabetes mellitus type 2, insulin resistance |
| HFpEF | hfpef, diastolic heart failure, preserved ejection fraction, heart failure with preserved ejection fraction, ef > 50, diastolic dysfunction, chf with preserved ef |
| HFrEF | hfref, systolic heart failure, reduced ejection fraction, heart failure with reduced ejection fraction, low ef, ef < 40, systolic dysfunction |
| Hypertension | hypertension, high blood pressure, htn, elevated bp, essential hypertension, chronic hypertension, uncontrolled hypertension |
| Metastatic Cancer | metastatic cancer, metastases, stage iv cancer, secondary cancer, cancer spread, distant metastasis, bone metastasis |
| Obesity | obesity, obese, morbid obesity, bmi > 30, bmi > 35, bmi > 40, severe obesity |
| Rheumatoid Arthritis | rheumatoid arthritis, ra, inflammatory arthritis, seropositive arthritis, seronegative arthritis, erosive arthritis, rheumatoid factor positive |
| SLE | sle, lupus, systemic lupus erythematosus, autoimmune lupus, lupus nephritis, lupus flare, lupus anticoagulant |
| VTE Complication | vte, dvt, pe, venous thromboembolism complication, deep vein thrombosis, pulmonary embolism, thromboembolism |
| Past VTE | history of vte, past dvt, previous pe, history of venous thromboembolism, prior dvt, prior pe, remote vte |

**Table S2:** Pre-consensus inter-annotator agreement metrics between physician and student annotators for multi-label annotation of discharge summaries (n = 1456).

| Metric | Value | Description |
|---|---|---|
| Mean Document Kappa ( ) | 0.805 | Average agreement across discharge summaries, calculated by treating each document's 17-label set as an observation. |
| Mean Label Kappa ( ) | 0.771 | Average agreement for individual clinical labels, calculated by comparing annotations for each label across all documents. |
| Total Unique Labels | 4542 | Union of all positive labels assigned by either annotator across all discharge summaries. |
| Total Disagreements | 1193 | Instances where physician and student annotations conflicted across all documents and labels. |
| Physician-Only Positives Disagreement | 927 | Labels assigned as positive by the physician but omitted by the student (potential over-labeling by physicians). |
| Student-Only Positives Disagreement | 266 | Labels assigned as positive by the student but omitted by the student (potential over-labeling by students). |

* Although 16 phenotypes, the 17th label was None for observations without any positive mention of the candidate phenotypes.

**Table S3:** Pre-consensus label-level inter-annotator agreement metrics (n = 1,456 discharge summaries)

| Label | | Total Disagreements | Disagreement Rate | Physician (+) / Student (−) | Physician (−) / Student (+) |
|---|---|---|---|---|---|
| Alcohol Abuse | 0.775 | 56 | 3.80% | 50 | 6 |
| C. difficile (complication) | 0.824 | 14 | 1.00% | 10 | 4 |
| C. difficile (medical history) | 0.921 | 15 | 1.00% | 0 | 15 |
| Dementia | 0.871 | 26 | 1.80% | 24 | 2 |
| Depression | 0.835 | 120 | 8.20% | 97 | 23 |
| Type I Diabetes | 0.799 | 51 | 3.50% | 20 | 31 |
| Type II Diabetes | 0.799 | 116 | 8.00% | 111 | 5 |
| Heart Failure (preserved EF) | 0.724 | 102 | 7.00% | 63 | 39 |
| Heart Failure (reduced EF) | 0.691 | 67 | 4.60% | 60 | 7 |
| Hypertension (essential) | 0.77 | 155 | 10.60% | 141 | 14 |
| Systemic Lupus | 0.964 | 8 | 0.50% | 2 | 6 |

| Label | | Total Disagreements | Disagreement Rate | Physician (+) / Student (−) | Physician (−) / Student (+) |
|---|---|---|---|---|---|
| Metastatic Cancer | 0.92 | 25 | 1.70% | 9 | 16 |
| Obesity | 0.227 | 272 | 18.70% | 256 | 16 |
| Rheumatoid Arthritis | 0.879 | 33 | 2.30% | 23 | 10 |
| DVT/PE (complication) | 0.727 | 23 | 1.60% | 11 | 12 |
| DVT/PE (medical history) | 0.877 | 60 | 4.10% | 47 | 13 |
| None | 0.505 | 50 | 3.40% | 3 | 47 |

Kappa ( ) scores and disagreement patterns for individual clinical concepts (n = 17 labels). Disagreement Rate = (Total Disagreements / 1,456) × 100.

Label-level agreement metrics demonstrate variability in annotator consensus. Disagreement rates reflect conflicts per 1,456 discharge summaries, with directional disagreements (Physician+/Student− vs. Physician−/Student+) revealing systematic biases. Obesity and 'None' labels show the highest discordance (see Discussion).

**Table S4:** Post-consensus Resolution of Annotator Disagreements

| Label | Total Disagreements | Rejected | Consensus | Agreed with Physician | Agreed with Student | Adjudicated Novel |
|---|---|---|---|---|---|---|
| Alcohol Abuse | 56 | 5 | 51 | 46 | 5 | 0 |
| C. difficile (complication) | 14 | 3 | 11 | 7 | 4 | 0 |
| C. difficile (medical history) | 15 | 4 | 11 | 0 | 11 | 0 |
| Dementia | 26 | 7 | 19 | 10 | 9 | 0 |
| Depression | 120 | 6 | 114 | 53 | 61 | 0 |
| Type I Diabetes | 51 | 16 | 35 | 31 | 4 | 0 |
| Type II Diabetes | 116 | 31 | 85 | 73 | 12 | 0 |
| Heart Failure (preserved EF) | 102 | 21 | 81 | 65 | 16 | 0 |
| Heart Failure (reduced EF) | 67 | 19 | 48 | 37 | 10 | 1 |
| Hypertension (essential) | 155 | 18 | 137 | 107 | 30 | 0 |
| Systemic Lupus | 8 | 0 | 8 | 5 | 3 | 0 |
| Metastatic Cancer | 25 | 4 | 21 | 14 | 7 | 0 |
| Obesity | 272 | 8 | 249 | 256 | 16 | 0 |
| Rheumatoid Arthritis | 33 | 0 | 33 | 22 | 11 | 0 |
| DVT/PE (complication) | 23 | 0 | 23 | 15 | 8 | 0 |
| DVT/PE (medical history) | 60 | 42 | 18 | 12 | 6 | 0 |
| None | 50 | 0 | 50 | 38 | 12 | 0 |
| Total | 1193 | 39 | 1154 | 760 | 394 | 1 |

Total disagreements reflect label-level conflicts between physician and medical student annotators. ”Rejected” indicates discharge summaries that did not undergo consensus review because of the predefined threshold of three or more disagreements per discharge summary. This explains why ”Rejected” column is not part of the total count of disagreements.

**Table S5:** Full results of EHR Phenotyping methods (baseline, supervised learning, large language model)

| Phenotype | Method | Model Configuration | F1 Score | Accuracy | Precision | Recall |
|---|---|---|---|---|---|---|
| AlcoholAb | ICD | ≥1 ICD code | 0.477 | 0.919 | 0.864 | 0.329 |
| AlcoholAb | ICD | ≥2 ICD codes | 0.050 | 0.891 | 1.000 | 0.026 |
| AlcoholAb | ICD | ≥3 ICD codes | 0.000 | 0.888 | 0.000 | 0.000 |
| AlcoholAb | TF-IDF | 25th percentile | 0.712 | 0.930 | 0.659 | 0.774 |
| AlcoholAb | TF-IDF | 50th percentile | 0.712 | 0.930 | 0.659 | 0.774 |
| AlcoholAb | TF-IDF | 75th percentile | 0.712 | 0.930 | 0.659 | 0.774 |
| AlcoholAb | TF-IDF | 90th percentile | 0.701 | 0.937 | 0.741 | 0.665 |
| AlcoholAb | Supervised | Lr | 0.614 | 0.914 | 0.627 | 0.603 |
| AlcoholAb | Supervised | Naive Bayes | 0.640 | 0.909 | 0.585 | 0.705 |
| AlcoholAb | Supervised | Random Forest | 0.369 | 0.796 | 0.285 | 0.526 |
| AlcoholAb | Supervised | Gradient Boosting | 0.229 | 0.578 | 0.145 | 0.551 |
| AlcoholAb | LLM | GPT-4o | 0.917 | 0.982 | 0.945 | 0.890 |
| CDiff_Comp | ICD | ≥1 ICD code | 0.860 | 0.991 | 0.784 | 0.952 |
| CDiff_Comp | ICD | ≥2 ICD codes | 0.000 | 0.970 | 0.000 | 0.000 |
| CDiff_Comp | ICD | ≥3 ICD codes | 0.000 | 0.970 | 0.000 | 0.000 |
| CDiff_Comp | TF-IDF | 25th percentile | 0.227 | 0.951 | 0.217 | 0.238 |
| CDiff_Comp | TF-IDF | 50th percentile | 0.227 | 0.951 | 0.217 | 0.238 |
| CDiff_Comp | TF-IDF | 75th percentile | 0.227 | 0.951 | 0.217 | 0.238 |
| CDiff_Comp | TF-IDF | 90th percentile | 0.227 | 0.951 | 0.217 | 0.238 |
| CDiff_Comp | Supervised | Lr | 0.057 | 0.036 | 0.029 | 0.952 |
| CDiff_Comp | Supervised | Naive Bayes | 0.114 | 0.886 | 0.075 | 0.238 |
| CDiff_Comp | Supervised | Random Forest | 0.442 | 0.930 | 0.292 | 0.905 |
| CDiff_Comp | Supervised | Gradient Boosting | 0.351 | 0.908 | 0.224 | 0.810 |
| CDiff_Comp | LLM | GPT-4o | 0.920 | 0.995 | 0.889 | 0.952 |
| CDiff_Hx | ICD | ≥1 ICD code | 0.368 | 0.931 | 0.549 | 0.277 |
| CDiff_Hx | ICD | ≥2 ICD codes | 0.000 | 0.927 | 0.000 | 0.000 |
| CDiff_Hx | ICD | ≥3 ICD codes | 0.000 | 0.927 | 0.000 | 0.000 |
| CDiff_Hx | TF-IDF | 25th percentile | 0.000 | 0.927 | 0.000 | 0.000 |
| CDiff_Hx | TF-IDF | 50th percentile | 0.000 | 0.927 | 0.000 | 0.000 |
| CDiff_Hx | TF-IDF | 75th percentile | 0.000 | 0.927 | 0.000 | 0.000 |
| CDiff_Hx | TF-IDF | 90th percentile | 0.000 | 0.927 | 0.000 | 0.000 |
| CDiff_Hx | Supervised | Lr | 0.137 | 0.151 | 0.074 | 0.920 |
| CDiff_Hx | Supervised | Naive Bayes | 0.218 | 0.864 | 0.188 | 0.260 |
| CDiff_Hx | Supervised | Random Forest | 0.425 | 0.905 | 0.381 | 0.480 |
| CDiff_Hx | Supervised | Gradient Boosting | 0.472 | 0.902 | 0.390 | 0.600 |
| CDiff_Hx | LLM | GPT-4o | 0.408 | 0.933 | 0.571 | 0.317 |
| DVT/PE_Comp | ICD | ≥1 ICD code | 0.236 | 0.921 | 0.162 | 0.436 |
| DVT/PE_Comp | ICD | ≥2 ICD codes | 0.360 | 0.977 | 0.818 | 0.231 |
| DVT/PE_Comp | ICD | ≥3 ICD codes | 0.095 | 0.973 | 0.667 | 0.051 |

| Phenotype | Method | Model Configuration | F1 Score | Accuracy | Precision | Recall |
|---|---|---|---|---|---|---|
| DVT/PE_Comp | TF-IDF | 25th percentile | 0.072 | 0.278 | 0.037 | 1.000 |
| DVT/PE_Comp | TF-IDF | 50th percentile | 0.085 | 0.517 | 0.045 | 0.795 |
| DVT/PE_Comp | TF-IDF | 75th percentile | 0.114 | 0.754 | 0.063 | 0.564 |
| DVT/PE_Comp | TF-IDF | 90th percentile | 0.112 | 0.886 | 0.072 | 0.256 |
| DVT/PE_Comp | Supervised | Lr | 0.000 | 0.971 | 0.000 | 0.000 |
| DVT/PE_Comp | Supervised | Naive Bayes | 0.122 | 0.895 | 0.081 | 0.250 |
| DVT/PE_Comp | Supervised | Random Forest | 0.194 | 0.806 | 0.110 | 0.800 |
| DVT/PE_Comp | Supervised | Gradient Boosting | 0.293 | 0.915 | 0.194 | 0.600 |
| DVT/PE_Comp | LLM | GPT-4o | 0.609 | 0.964 | 0.438 | 1.000 |
| DVT/PE_Hx | ICD | ≥1 ICD code | 0.149 | 0.744 | 0.295 | 0.099 |
| DVT/PE_Hx | ICD | ≥2 ICD codes | 0.019 | 0.772 | 0.273 | 0.010 |
| DVT/PE_Hx | ICD | ≥3 ICD codes | 0.006 | 0.774 | 0.333 | 0.003 |
| DVT/PE_Hx | TF-IDF | 25th percentile | 0.000 | 0.775 | 0.000 | 0.000 |
| DVT/PE_Hx | TF-IDF | 50th percentile | 0.000 | 0.775 | 0.000 | 0.000 |
| DVT/PE_Hx | TF-IDF | 75th percentile | 0.000 | 0.775 | 0.000 | 0.000 |
| DVT/PE_Hx | TF-IDF | 90th percentile | 0.000 | 0.775 | 0.000 | 0.000 |
| DVT/PE_Hx | Supervised | Lr | 0.345 | 0.561 | 0.260 | 0.513 |
| DVT/PE_Hx | Supervised | Naive Bayes | 0.312 | 0.711 | 0.336 | 0.292 |
| DVT/PE_Hx | Supervised | Random Forest | 0.351 | 0.637 | 0.294 | 0.435 |
| DVT/PE_Hx | Supervised | Gradient Boosting | 0.349 | 0.678 | 0.321 | 0.383 |
| DVT/PE_Hx | LLM | GPT-4o | 0.871 | 0.934 | 0.775 | 0.994 |
| Dementia | ICD | ≥1 ICD code | 0.603 | 0.955 | 0.870 | 0.461 |
| Dementia | ICD | ≥2 ICD codes | 0.019 | 0.927 | 1.000 | 0.010 |
| Dementia | ICD | ≥3 ICD codes | 0.000 | 0.927 | 0.000 | 0.000 |
| Dementia | TF-IDF | 25th percentile | 0.467 | 0.839 | 0.308 | 0.961 |
| Dementia | TF-IDF | 50th percentile | 0.467 | 0.839 | 0.308 | 0.961 |
| Dementia | TF-IDF | 75th percentile | 0.467 | 0.839 | 0.308 | 0.961 |
| Dementia | TF-IDF | 90th percentile | 0.672 | 0.943 | 0.583 | 0.794 |
| Dementia | Supervised | Lr | 0.176 | 0.673 | 0.105 | 0.545 |
| Dementia | Supervised | Naive Bayes | 0.348 | 0.874 | 0.261 | 0.523 |
| Dementia | Supervised | Random Forest | 0.466 | 0.896 | 0.348 | 0.705 |
| Dementia | Supervised | Gradient Boosting | 0.476 | 0.887 | 0.340 | 0.795 |
| Dementia | LLM | GPT-4o | 0.896 | 0.983 | 0.832 | 0.971 |
| Depression | ICD | ≥1 ICD code | 0.799 | 0.830 | 0.947 | 0.691 |
| Depression | ICD | ≥2 ICD codes | 0.003 | 0.512 | 1.000 | 0.001 |
| Depression | ICD | ≥3 ICD codes | 0.000 | 0.511 | 0.000 | 0.000 |
| Depression | TF-IDF | 25th percentile | 0.916 | 0.919 | 0.921 | 0.912 |
| Depression | TF-IDF | 50th percentile | 0.916 | 0.919 | 0.921 | 0.912 |
| Depression | TF-IDF | 75th percentile | 0.622 | 0.720 | 0.919 | 0.470 |
| Depression | TF-IDF | 90th percentile | 0.313 | 0.595 | 0.921 | 0.189 |
| Depression | Supervised | Lr | 0.419 | 0.555 | 0.579 | 0.328 |
| Depression | Supervised | Naive Bayes | 0.524 | 0.620 | 0.678 | 0.427 |

| Phenotype | Method | Model Configuration | F1 Score | Accuracy | Precision | Recall |
|---|---|---|---|---|---|---|
| Depression | Supervised | Random Forest | 0.685 | 0.616 | 0.572 | 0.854 |
| Depression | Supervised | Gradient Boosting | 0.675 | 0.644 | 0.610 | 0.755 |
| Depression | LLM | GPT-4o | 0.894 | 0.891 | 0.855 | 0.937 |
| HFpEF | ICD | ≥1 ICD code | 0.853 | 0.955 | 0.863 | 0.843 |
| HFpEF | ICD | ≥2 ICD codes | 0.000 | 0.844 | 0.000 | 0.000 |
| HFpEF | ICD | ≥3 ICD codes | 0.000 | 0.844 | 0.000 | 0.000 |
| HFpEF | TF-IDF | 25th percentile | 0.423 | 0.884 | 0.952 | 0.272 |
| HFpEF | TF-IDF | 50th percentile | 0.423 | 0.884 | 0.952 | 0.272 |
| HFpEF | TF-IDF | 75th percentile | 0.423 | 0.884 | 0.952 | 0.272 |
| HFpEF | TF-IDF | 90th percentile | 0.423 | 0.884 | 0.952 | 0.272 |
| HFpEF | Supervised | Lr | 0.345 | 0.540 | 0.223 | 0.769 |
| HFpEF | Supervised | Naive Bayes | 0.426 | 0.796 | 0.382 | 0.481 |
| HFpEF | Supervised | Random Forest | 0.682 | 0.879 | 0.582 | 0.824 |
| HFpEF | Supervised | Gradient Boosting | 0.731 | 0.907 | 0.669 | 0.806 |
| HFpEF | LLM | GPT-4o | 0.847 | 0.948 | 0.784 | 0.922 |
| HFrEF | ICD | ≥1 ICD code | 0.810 | 0.967 | 0.817 | 0.803 |
| HFrEF | ICD | ≥2 ICD codes | 0.032 | 0.913 | 1.000 | 0.016 |
| HFrEF | ICD | ≥3 ICD codes | 0.000 | 0.912 | 0.000 | 0.000 |
| HFrEF | TF-IDF | 25th percentile | 0.351 | 0.928 | 0.844 | 0.221 |
| HFrEF | TF-IDF | 50th percentile | 0.351 | 0.928 | 0.844 | 0.221 |
| HFrEF | TF-IDF | 75th percentile | 0.351 | 0.928 | 0.844 | 0.221 |
| HFrEF | TF-IDF | 90th percentile | 0.351 | 0.928 | 0.844 | 0.221 |
| HFrEF | Supervised | Lr | 0.167 | 0.127 | 0.091 | 1.000 |
| HFrEF | Supervised | Naive Bayes | 0.408 | 0.868 | 0.337 | 0.517 |
| HFrEF | Supervised | Random Forest | 0.325 | 0.763 | 0.217 | 0.650 |
| HFrEF | Supervised | Gradient Boosting | 0.293 | 0.640 | 0.177 | 0.850 |
| HFrEF | LLM | GPT-4o | 0.865 | 0.974 | 0.799 | 0.943 |
| HTN | ICD | ≥1 ICD code | 0.668 | 0.648 | 0.926 | 0.522 |
| HTN | ICD | ≥2 ICD codes | 0.000 | 0.323 | 0.000 | 0.000 |
| HTN | ICD | ≥3 ICD codes | 0.000 | 0.323 | 0.000 | 0.000 |
| HTN | TF-IDF | 25th percentile | 0.899 | 0.855 | 0.855 | 0.947 |
| HTN | TF-IDF | 50th percentile | 0.776 | 0.736 | 0.914 | 0.674 |
| HTN | TF-IDF | 75th percentile | 0.519 | 0.554 | 0.963 | 0.355 |
| HTN | TF-IDF | 90th percentile | 0.250 | 0.417 | 0.971 | 0.144 |
| HTN | Supervised | Lr | 0.607 | 0.604 | 0.921 | 0.452 |
| HTN | Supervised | Naive Bayes | 0.778 | 0.727 | 0.861 | 0.710 |
| HTN | Supervised | Random Forest | 0.491 | 0.526 | 0.897 | 0.338 |
| HTN | Supervised | Gradient Boosting | 0.491 | 0.526 | 0.897 | 0.338 |
| HTN | LLM | GPT-4o | 0.948 | 0.927 | 0.907 | 0.994 |
| MetCancer | ICD | ≥1 ICD code | 0.892 | 0.974 | 0.898 | 0.887 |
| MetCancer | ICD | ≥2 ICD codes | 0.730 | 0.947 | 0.943 | 0.595 |
| MetCancer | ICD | ≥3 ICD codes | 0.459 | 0.914 | 0.944 | 0.304 |
| MetCancer | TF-IDF | 25th percentile | 0.701 | 0.937 | 0.829 | 0.607 |

| Phenotype | Method | Model Configuration | F1 Score | Accuracy | Precision | Recall |
|---|---|---|---|---|---|---|
| MetCancer | TF-IDF | 50th percentile | 0.701 | 0.937 | 0.829 | 0.607 |
| MetCancer | TF-IDF | 75th percentile | 0.701 | 0.937 | 0.829 | 0.607 |
| MetCancer | TF-IDF | 90th percentile | 0.701 | 0.937 | 0.829 | 0.607 |
| MetCancer | Supervised | Lr | 0.253 | 0.474 | 0.153 | 0.726 |
| MetCancer | Supervised | Naive Bayes | 0.552 | 0.886 | 0.533 | 0.571 |
| MetCancer | Supervised | Random Forest | 0.544 | 0.809 | 0.384 | 0.929 |
| MetCancer | Supervised | Gradient Boosting | 0.564 | 0.826 | 0.407 | 0.917 |
| MetCancer | LLM | GPT-4o | 0.900 | 0.973 | 0.826 | 0.988 |
| Obesity | ICD | ≥1 ICD code | 0.736 | 0.901 | 0.814 | 0.671 |
| Obesity | ICD | ≥2 ICD codes | 0.021 | 0.796 | 1.000 | 0.010 |
| Obesity | ICD | ≥3 ICD codes | 0.000 | 0.794 | 0.000 | 0.000 |
| Obesity | TF-IDF | 25th percentile | 0.776 | 0.890 | 0.670 | 0.923 |
| Obesity | TF-IDF | 50th percentile | 0.776 | 0.890 | 0.670 | 0.923 |
| Obesity | TF-IDF | 75th percentile | 0.793 | 0.906 | 0.723 | 0.878 |
| Obesity | TF-IDF | 90th percentile | 0.607 | 0.880 | 0.928 | 0.451 |
| Obesity | Supervised | Lr | 0.326 | 0.632 | 0.264 | 0.427 |
| Obesity | Supervised | Naive Bayes | 0.299 | 0.720 | 0.313 | 0.287 |
| Obesity | Supervised | Random Forest | 0.405 | 0.606 | 0.296 | 0.643 |
| Obesity | Supervised | Gradient Boosting | 0.415 | 0.527 | 0.280 | 0.804 |
| Obesity | LLM | GPT-4o | 0.821 | 0.913 | 0.712 | 0.969 |
| RA | ICD | ≥1 ICD code | 0.751 | 0.947 | 0.853 | 0.671 |
| RA | ICD | ≥2 ICD codes | 0.024 | 0.883 | 1.000 | 0.012 |
| RA | ICD | ≥3 ICD codes | 0.012 | 0.883 | 1.000 | 0.006 |
| RA | TF-IDF | 25th percentile | 0.224 | 0.326 | 0.130 | 0.823 |
| RA | TF-IDF | 50th percentile | 0.219 | 0.517 | 0.135 | 0.573 |
| RA | TF-IDF | 75th percentile | 0.145 | 0.685 | 0.107 | 0.226 |
| RA | TF-IDF | 90th percentile | 0.086 | 0.800 | 0.094 | 0.079 |
| RA | Supervised | Lr | 0.237 | 0.689 | 0.168 | 0.402 |
| RA | Supervised | Naive Bayes | 0.406 | 0.838 | 0.362 | 0.463 |
| RA | Supervised | Random Forest | 0.723 | 0.923 | 0.633 | 0.841 |
| RA | Supervised | Gradient Boosting | 0.349 | 0.678 | 0.321 | 0.383 |
| RA | LLM | GPT-4o | 0.920 | 0.980 | 0.870 | 0.976 |
| SLE | ICD | ≥1 ICD code | 0.789 | 0.960 | 0.686 | 0.929 |
| SLE | ICD | ≥2 ICD codes | 0.144 | 0.923 | 0.750 | 0.080 |
| SLE | ICD | ≥3 ICD codes | 0.018 | 0.919 | 1.000 | 0.009 |
| SLE | TF-IDF | 25th percentile | 0.343 | 0.688 | 0.207 | 1.000 |
| SLE | TF-IDF | 50th percentile | 0.343 | 0.688 | 0.207 | 1.000 |
| SLE | TF-IDF | 75th percentile | 0.483 | 0.829 | 0.320 | 0.982 |
| SLE | TF-IDF | 90th percentile | 0.706 | 0.947 | 0.640 | 0.788 |
| SLE | Supervised | Lr | 0.313 | 0.917 | 0.481 | 0.232 |
| SLE | Supervised | Naive Bayes | 0.556 | 0.914 | 0.481 | 0.661 |
| SLE | Supervised | Random Forest | 0.442 | 0.804 | 0.288 | 0.946 |

| Phenotype | Method | Model Configuration | F1 Score | Accuracy | Precision | Recall |
|---|---|---|---|---|---|---|
| SLE | Supervised | Gradient Boosting | 0.781 | 0.959 | 0.694 | 0.893 |
| SLE | LLM | GPT-4o | 0.953 | 0.992 | 0.918 | 0.991 |
| T1D | ICD | ≥1 ICD code | 0.535 | 0.937 | 0.962 | 0.370 |
| T1D | ICD | ≥2 ICD codes | 0.469 | 0.932 | 0.955 | 0.311 |
| T1D | ICD | ≥3 ICD codes | 0.379 | 0.924 | 0.941 | 0.237 |
| T1D | TF-IDF | 25th percentile | 0.452 | 0.930 | 0.952 | 0.296 |
| T1D | TF-IDF | 50th percentile | 0.452 | 0.930 | 0.952 | 0.296 |
| T1D | TF-IDF | 75th percentile | 0.452 | 0.930 | 0.952 | 0.296 |
| T1D | TF-IDF | 90th percentile | 0.452 | 0.930 | 0.952 | 0.296 |
| T1D | Supervised | Lr | 0.460 | 0.863 | 0.377 | 0.588 |
| T1D | Supervised | Naive Bayes | 0.460 | 0.863 | 0.377 | 0.588 |
| T1D | Supervised | Random Forest | 0.529 | 0.857 | 0.393 | 0.809 |
| T1D | Supervised | Gradient Boosting | 0.471 | 0.803 | 0.321 | 0.882 |
| T1D | LLM | GPT-4o | 0.923 | 0.985 | 0.913 | 0.933 |
| T2D | ICD | ≥1 ICD code | 0.853 | 0.906 | 0.764 | 0.964 |
| T2D | ICD | ≥2 ICD codes | 0.263 | 0.726 | 0.548 | 0.173 |
| T2D | ICD | ≥3 ICD codes | 0.148 | 0.719 | 0.523 | 0.087 |
| T2D | TF-IDF | 25th percentile | 0.342 | 0.739 | 0.599 | 0.239 |
| T2D | TF-IDF | 50th percentile | 0.342 | 0.739 | 0.599 | 0.239 |
| T2D | TF-IDF | 75th percentile | 0.342 | 0.739 | 0.599 | 0.239 |
| T2D | TF-IDF | 90th percentile | 0.327 | 0.742 | 0.626 | 0.221 |
| T2D | Supervised | Lr | 0.657 | 0.785 | 0.600 | 0.727 |
| T2D | Supervised | Naive Bayes | 0.460 | 0.863 | 0.377 | 0.588 |
| T2D | Supervised | Random Forest | 0.752 | 0.829 | 0.639 | 0.912 |
| T2D | Supervised | Gradient Boosting | 0.693 | 0.776 | 0.567 | 0.892 |
| T2D | LLM | GPT-4o | 0.939 | 0.964 | 0.902 | 0.980 |
